# Genetic relationships and causality between overall and central adiposity and breast, prostate, lung and colorectal cancer

**DOI:** 10.1101/2022.12.19.22283607

**Authors:** Jared G Maina, Vincent Pascat, Liudmila Zudina, Anna Ulrich, Igor Pupko, Ayse Demirkan, Amélie Bonnefond, Zhanna Balkhiyarova, Marika Kaakinen, Philippe Froguel, Inga Prokopenko

**Affiliations:** INSERM UMR 1283, CNRS UMR 8199, European Genomic Institute for Diabetes (EGID), Institut Pasteur de Lille, F-59000 Lille, France; University of Lille, Lille University Hospital, Lille, F-59000, France; Section of Statistical Multi-Omics, Department of Clinical and Experimental Medicine, University of Surrey, Guildford, United Kingdom; Department of Metabolism Digestion and Reproduction, Imperial College London, London United Kingdom; People-Centred Artificial Intelligence Institute, University of Surrey, Guildford, United Kingdom

**Keywords:** Obesity, Cancer, Mendelian randomization, polygenic scores

## Abstract

**OBJECTIVE:** Diverse measures of obesity relate to cancer risk differently. Here we assess the relationship between overall and central adiposity and cancer.

**METHODS:** We constructed z-score weighted polygenic scores (PGS) for two obesity-related phenotypes; body mass index (BMI) and BMI adjusted waist-to-hip ratio (WHRadjBMI) and tested for their association with five cancers in the UK Biobank: overall breast (BrC), post-menopausal breast (PostBrC), prostate (PrC), colorectal (CrC) and lung (LungC) cancer. We utilised publicly available data to perform bi-directional Mendelian randomization (MR) between BMI/WHRadjBMI and BrC, PrC and CrC.

**RESULTS:** PGS_BMI_ had significant multiple testing-corrected inverse association with PrC (OR[95%CI]=0.97[0.95-0.99], *P*=0.0012) but PGS_WHRadjBMI_ was not associated with PrC. PGS_BMI_ was associated with PostBrC (OR[95%CI]=0.97[0.96-0.99], *P*=0.00203) while PGS_WHRadjBMI_ had nominal association with BrC. PGS_BMI_ had nominal positive association with LungC. MR analyses showed significant multiple testing-corrected protective causal effect of BMI on PrC (OR[95%CI]=0.993[0.988-0.998], *P*=4.19×10^−3^). WHRadjBMI had a nominal causal effect on higher PrC risk (OR[95%CI]=1.022[1.0067-1.038], *P*=0.0053). We also report nominal causal protective effect of WHRadjBMI on breast cancer (OR[95%CI]=0.99[0.98-0.997], *P*=0.0068). Neither PGS nor MR analyses were significant for CrC.

**CONCLUSIONS:** Higher overall adiposity appears protective from PrC while higher central adiposity is a potential risk factor for PrC but protective from BrC.

**STUDY IMPORTANCE:** *What is already known about this subject?:*

- Observational studies suggest obesity is associated with higher risk of certain cancers and at the same time is protective of other cancers. The direction of association is in part influenced by the anthropometric trait used to assess obesity.
- Higher BMI appears protective from prostate, breast and lung cancers but is a risk factor for post-menopausal breast, pancreatic and colorectal cancers.

*What are the new findings in your manuscript?:*

- We implement Mendelian randomization approach using large scale datasets and show a protective causal effect of higher BMI from prostate cancer but suggest that higher WHRadjBMI is causal for prostate cancer.
- We also show nominal evidence of WHRadjBMI being causally protective from breast cancer.

*How might your results change the direction of research or the focus of clinical practice?:*

- We demonstrate the importance of partitioning obesity into discrete types depending on the area of fat deposition rather than using an overall measure.
- Our results show that diverse measures of obesity relate differently to cancer risk. In fact, even for the same type of cancer, overall and central obesity measures may impact in opposite direction in terms of risk to cancer.

## INTRODUCTION

Two common anthropometric measures used to define obesity are body mass index (BMI) and waist-to-hip circumference ratio (WHR). They represent the body’s overall and central abdominal adiposity, respectively. Despite being a routine measure of adiposity, BMI does not accurately capture well body composition as it does not distinguish lean mass from fat mass(1). Additionally, individuals who are within normal BMI ranges may be metabolically unhealthy(2), and others with elevated BMI present normal metabolic parameters. Other measures of adiposity have hence been used to improve clinical evaluations of metabolic health, including WHR. High WHR is associated with insulin resistance and contributes to the definition of the metabolic syndrome(3). Epidemiological evidence suggests BMI is positively associated with increased cancer risk of post-menopausal breast cancer(4), pancreatic cancer(5), colorectal cancer(6), while inversely associated with prostate cancer(7), pre-menopausal breast cancer and lung and oral cavity cancers(4). The relationship between cancer incidence, its mortality, and WHR is relatively understudied compared to that with BMI. Despite this, WHR appears to be a better predictor of cancer risk compared to BMI(8,9). However, correlation observed in epidemiological studies does not infer causality. Moreover, observational studies are prone to bias by unadjusted confounders such as tobacco use in lung cancer studies(4). To date, genome-wide association studies (GWAS) have identified hundreds of genomic loci, associated with BMI and body fat(10,11) and many loci with WHR adjusted for BMI (WHRadjBMI)(11) with a relatively small overlap between the two sets of associated loci. GWAS have also found many loci associated with cancer phenotypes(12–15). However, one limitation of GWAS is that these loci identified individually for adiposity and cancer phenotypes offer little evidence of the shared pathophysiology between them. Despite the above-mentioned limitation of GWAS, genomic loci identified in GWAS can be implemented in methods such as polygenic scores (PGS)(16) and Mendelian randomization (MR)(17). PGS refer to the weighted (by effect size) sum of risk variants identified from GWAS that an individual has for a particular phenotype(16). PGS are useful for risk prediction analyses as well as association testing. In MR analyses, genetic variants identified in GWAS, referred to as instrument variables, are used to estimate the causal effect of a risk factor (exposure) on an outcome of interest(17).

In this study, we aimed to assess the impact of overall and central adiposity on cancer risk. First, we defined the genetic correlation between BMI/WHRadjBMI and cancers using the UK Biobank (UKBB) dataset. Additionally, we used established BMI and WHRadjBMI genome-wide loci(10,11) (*P*<5×10^−8^) to create PGS which were then tested for association with five cancer phenotypes in UKBB including overall breast, post-menopausal breast, colorectal, prostate and lung cancer. Further, using established genetic variants, associated with these phenotypes, we performed MR between the two adiposity traits and three cancers (breast, prostate and colorectal) to investigate the causal relationships between them. The MR analyses did not include post-menopausal breast and lung cancer due to the unavailability of summary statistics.

## METHODS

### UK Biobank

We used the UKBB resource (www.ukbiobank.ac.uk) to define adiposity and cancer phenotypes. The UKBB includes approximately 500,000 individuals, aged between 40-69 years recruited from 22 centres across the United Kingdom. Phenotypic data collected from recruited participants includes biological samples, physical measurements and responses from a questionnaire administered at recruitment. Genetic data is available for 488,377 individuals who were genotyped using the UKBB BiLEVE array (n=49,979) and the UK Biobank Axiom Array (n=438,398)(18). 457,270 individuals of European ancestry had weight (kg), height (m), waist and hip circumference (cm) measurements which were used to define the BMI (weight/[height]^2^ and WHR (waist/hip) phenotypes. Cancer phenotypes were defined using the criteria described in **Table S1**. There were 18,676 overall breast cancer, 13,355 postmenopausal breast cancer, 11,825 prostate cancer, 8,201 colorectal cancer and 4,237 lung cancer cases (**Table S2**).

### Genetic correlation and SNP heritability estimation

We used the LD Score (LDSC) regression approach and tool(19) to estimate the genetic correlation (rG) between adiposity phenotypes and cancer in the UKBB. The proportion of genetic variance explained by genome-wide SNPs (h^2^_SNP_) for each of our UKBB phenotypes was also computed using the LDSC tool. UKBB GWAS summary statistics were filtered based on the following parameters: imputation score> 0.9, minor allele frequency (MAF) > 0.01 and 0.1 ≥ *P* > 0. Ambiguous strand, duplicated SNPs and variants that did not represent SNPs (e.g., indels) were removed. The Bonferroni corrected significance threshold to determine significant genetic correlation estimates was set as P<0.005 (0.05/10, the number of genetic correlation tests done in our analyses; five for BMI and five for WHRadjBMI). Nominal significance threshold was set at 0.05 ≥ *P* > 0.005.

### Polygenic scores

We used the adiposity-increasing alleles at genome-wide significant variants from recent large scale meta-analyses of BMI (670 SNPs) and WHRadjBMI (346 SNPs) to construct PGSs. The SNP sets for BMI and WHRadjBMI PGSs base data were derived from the latest adiposity GWAS meta-analyses from the GIANT consortium(10,11). These studies meta-analysed UKBB with previous BMI and WHRadjBMI studies(20,21). Since the target data (UKBB) was part of the adiposity meta-analyses which formed the PGS base data in this analysis, we used variant effect size estimates from earlier studies which has not included UKBB in the meta-analyses for the weighted PGS(20,21). In total, there 567 and 274 SNPs for BMI and WHRadjBMI PGS analyses (**Figure 1**). The PGSs were generated using PLINK version 1.90b4.1(22). The PGSs were tested for association with five cancers in UKBB using logistic regression models in RStudio(23). For sex-specific cancers (breast and prostate), we included age, batch array and six principal components (PCs) as covariates in the regression model. For all the other cancers, sex was included as an additional covariate. Additionally, we performed BMI PGS association tests by splitting UKBB dataset into four BMI categories for each of the cancers (underweight [BMI < 18.5kg/m^2^], normal weight [25kg/m^2^ ≥ BMI > 18.5kg/m^2^], overweight [30kg/m^2^ ≥ BMI > 25kg/m^2^], obese [BMI > 30kg/m^2^]). The Bonferroni corrected significance threshold (Bonferroni^PGS^) to determine significant associations was set as P<0.005 (0.05/10, the number of association tests done in our analyses; five for BMI and five for WHRadjBMI). Nominal significance threshold was set at 0.05 ≥ P > 0.005.

**Figure 1.**
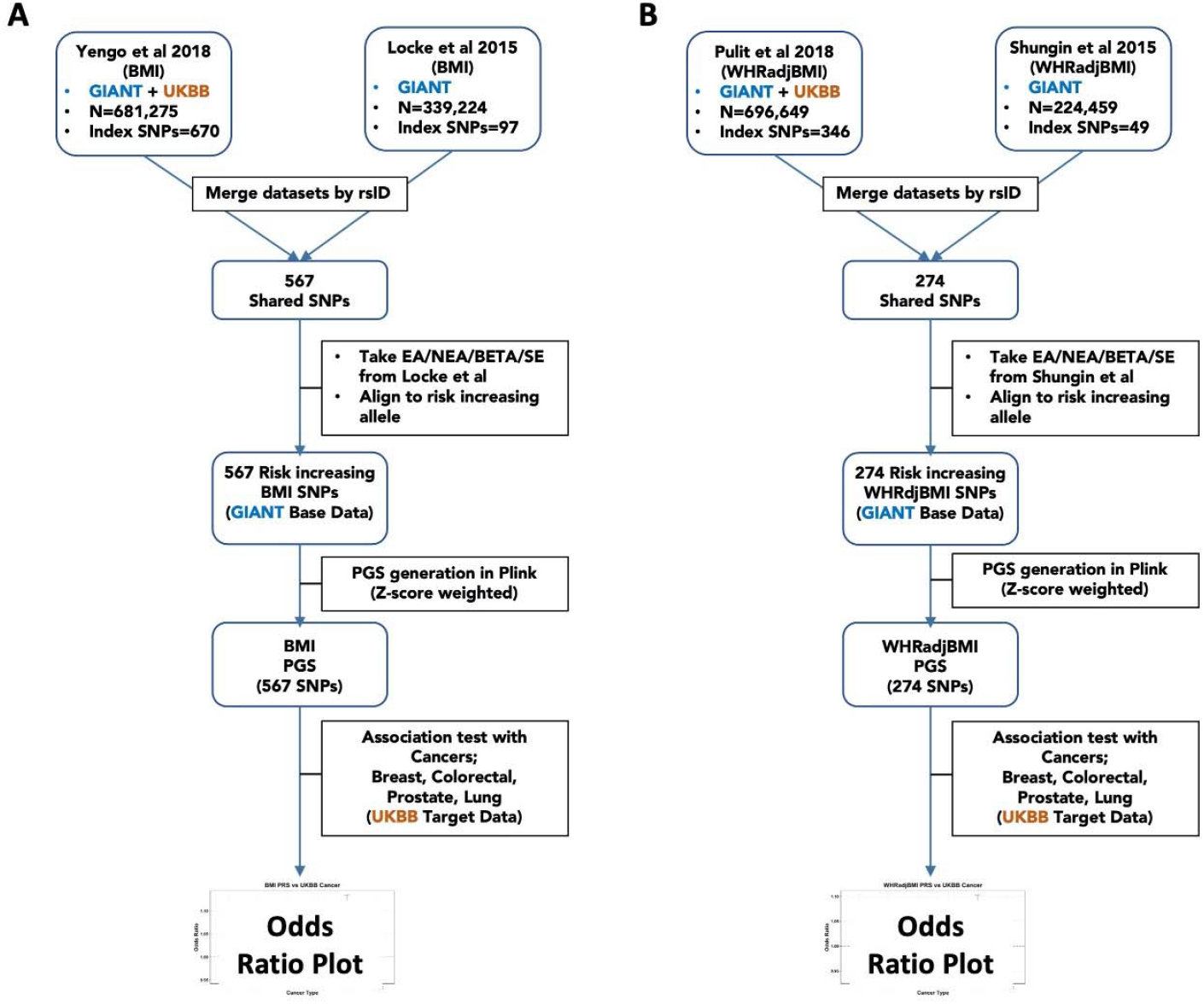
BMI (A) and WHRadjBMI (B) Polygenic score analyses pipeline. Legend: A two-sample approach was used to construct our PGS base data. The SNPs used for the PGS were from GIANT’s latest BMI and WHRadjBMI meta-analyses. Since the meta-analyses comprised of the UK Biobank (our target data), we used weights from the non-UK Biobank study in GIANT’s meta-analyses

### Mendelian Randomization

To investigate the causality between BMI/WHRadjBMI and cancer, we performed bi-directional MR using the *TwoSampleMR* R package version 0.5.6(24). We tested the effect of adiposity (BMI and WHRadjBMI) as an exposure for cancer outcomes, and the reverse direction with cancers as risk factors to adiposity (BMI and WHRadjBMI) using summary statistics from independent datasets. Independent, genome-wide significant SNPs (*P*<5×10^−8^) were used as genetic instruments for MR in our analyses. We obtained the genetic instruments for BMI (670 SNPs) and WHRadjBMI (346 SNPs) from the GIANT consortium(10,11). Furthermore, the cancer genetic instruments were derived from recent large-scale GWAS of breast (201 SNPs)(12), colorectal (137 SNPs)(14) and prostate (248 SNPs)(13) cancers using genome-wide significant SNPs. The number of SNPs available for each MR test are summarised in **Figure S1**. Moreover, the causal effect estimate was obtained using the inverse-variance weighted (MR-IVW) method(25), which combines the ratio estimates of individual variants using a random-effect meta-analysis. Sensitivity MR analyses were performed using MR Egger, weighted median, weighted mode and simple mode methods(26). Exclusion of palindromic SNPs from the exposure-outcome pairs, as well as allele matching was performed as part of the *TwoSampleMR* pipeline. Additional quality control steps included removal of outliers after inspection of scatter plots and leave-one-out results also performed using the MR package. We also performed the Cochran’s Q test as part of the MR pipeline to assess heterogeneity on our instruments. The Bonferroni corrected significance threshold (Bonferroni^MR^) to determine significant associations was set as *P*<0.0042 (0.05/12, the number of MR tests done in our analyses; **Figure S1**). Nominal significance threshold was set at 0.05 ≥ *P* > 0.00442.

## RESULTS

### Definition of overlap of the loci between adiposity and cancer phenotypes in the UKBB

We assessed the overlap of genome-wide significant loci between the cancers and adiposity phenotypes using the UKBB GWAS summary statistics (**Supplementary data**). There was no loci overlap between adiposity phenotypes and either colorectal and lung cancers. However, we observed overlap between the sex-specific cancers (BrC, PostBrC and PrC) and both BMI and WHRadjBMI. There were five shared loci between BMI and BrC in UKBB (**Figure 2a**): *FTO, EBF1, ERBB4, TBX3/MED13L* and *CASC16*. In contrast, there was no overlap between PostBrC and BMI. Furthermore, three loci were shared between BMI and PrC (**Figure 2a**): *TMEM17/EHBP1, JAZF1* and *MIR4686/ASCL2*. In relation to WHRadjBMI, eight loci were shared between BrC (**Figure 2b**): *ZMIZ1, NRIP1/USP25, EBF1, ESR1, RAD51B, CASC21/CASC8, CCND1* and *FGFR2*. Five loci were shared between PostBrC and WHRadjBMI (*EBF1, ESR1, RAD51B, CCND1, CASC21/CASC8*).

**Figure 2.**
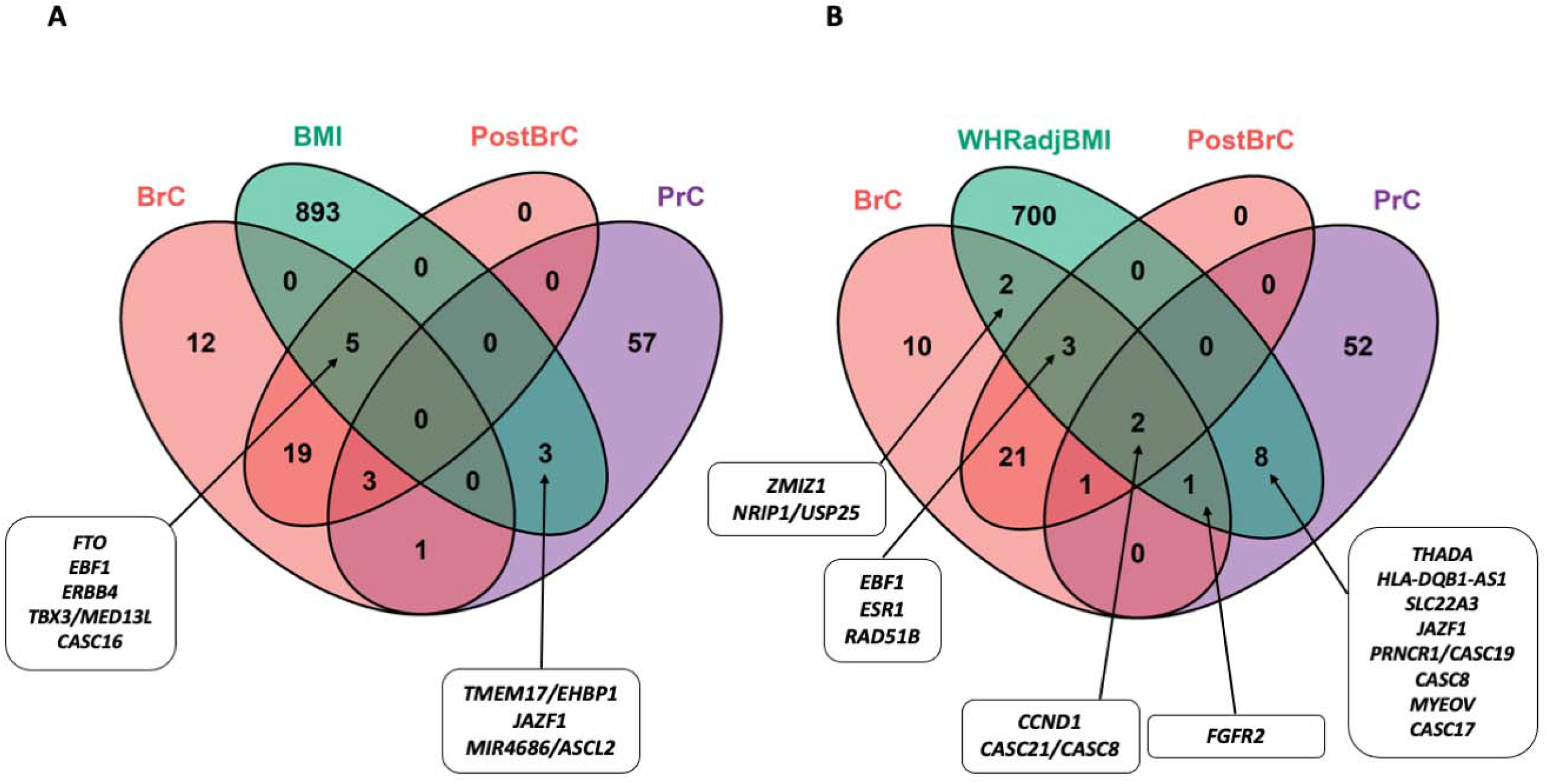
Venn diagram defining overlapping genome-wide significant loci between adiposity phenotypes and sex-specific cancers based on UK Biobank GWAS summary statistics. Legend: A) Loci overlap between BMI and the sex-specific cancers in the UK Biobank. B) Loci overlap between WHRadjBMI and the sex-specific cancers in the UK Biobank. BrC= overall breast cancer, PostBrC=post-menopausal breast cancer, PrC=prostate cancer.

Further, 11 loci were shared between PrC and WHRadjBMI (**Figure 2b**): *THADA, HLA-DQB1-AS1, SLC22A3, JAZF1, PRNCR1/CASC19, CASC8, MYEOV, CASC17, FGFR2, CCND1, CASC21/CASC8*.

### Genetic correlation and SNP heritability estimates

The UKBB genetic correlation analyses identified nominally significant inverse genetic correlation between BMI and prostate cancer (rG=-0.076, *P*=0.0075). Both BMI and WHRadjBMI had a nominally significant positive genetic correlation with lung cancer in UKBB (rG_BMI_=0.18, *P*=0.0014, rG_WHRadjBMI_=0.16, *P*=0.0065). Nominal inverse genetic correlation was also observed between BMI and post-menopausal breast cancer (rG=-0.0803, *P*=0.014), whereas positive genetic correlation was observed between WHRadjBMI and colorectal cancer (rG=0.103, *P*=0.017) (**Figure 3a, Table S3**).

**Figure 3.**
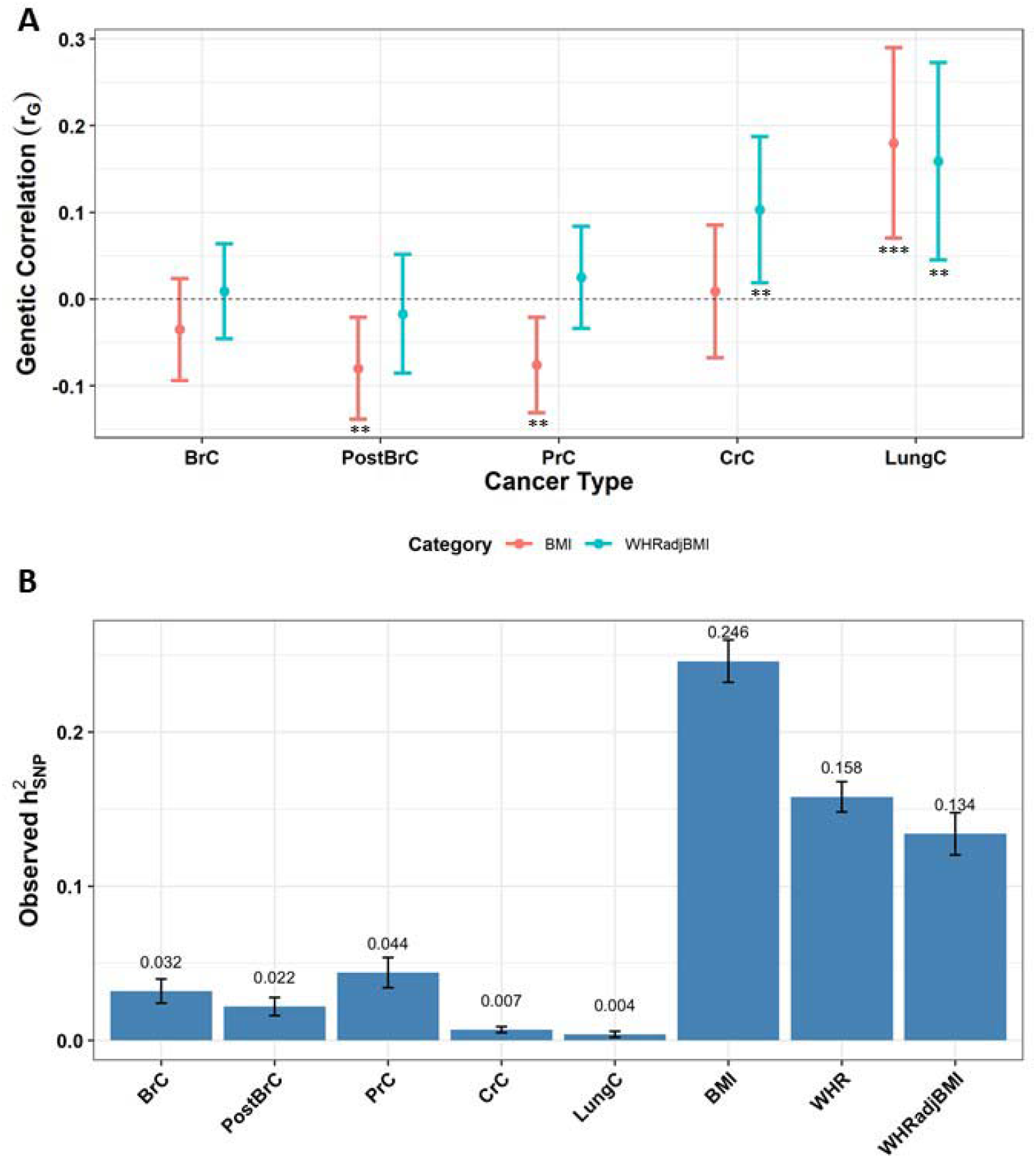
Genetic correlation and SNP heritability estimates of BMI/WHRadjBMI and cancer in UK Biobank. Legend: A). Genetic correlation estimates between UK Biobank BMI/WHRadjBMI and cancer. Estimates which withstood Bonferroni corrections (*P*<0.05/10=0.005) are marked with triple asterisks (***), double asterisks (**) for 0.05 ≥ *P* > 0.005. BrC=overall breast cancer, PostBrC=post-menopausal breast cancer, PrC=prostate cancer, CrC=colorectal cancer, LungC=lung cancer. B). SNP heritability estimates of BMI/WHRadjBMI and cancer in UK Biobank

The observed SNP heritability estimates (h^2^_SNP_) for the BMI and WHRadjBMI were 24.59% and 13.43% respectively. Estimates for cancer ranged from 0.36% (lung) to 4.41% (prostate) (**Figure 3b** and **Table S4**).

### Association analyses with polygenic scores

BMI PGS (567 SNPs) had a significant, after correction for multiple testing, inverse association with prostate (OR[95%CI]=0.97[0.95-0.99], *P*=0.0012) and post-menopausal breast (OR[95%CI]=0.97[0.96-0.99], *P*=0.00203) cancers (**Figure 4, Table 1**). We also identified nominal associations with overall breast cancer (OR[95%CI]=0.98[0.96-0.99], *P*=0.0086) and lung cancer (OR[95%CI]=1.044[1.013-1.076], *P*=0.0057).

**Figure 4.**
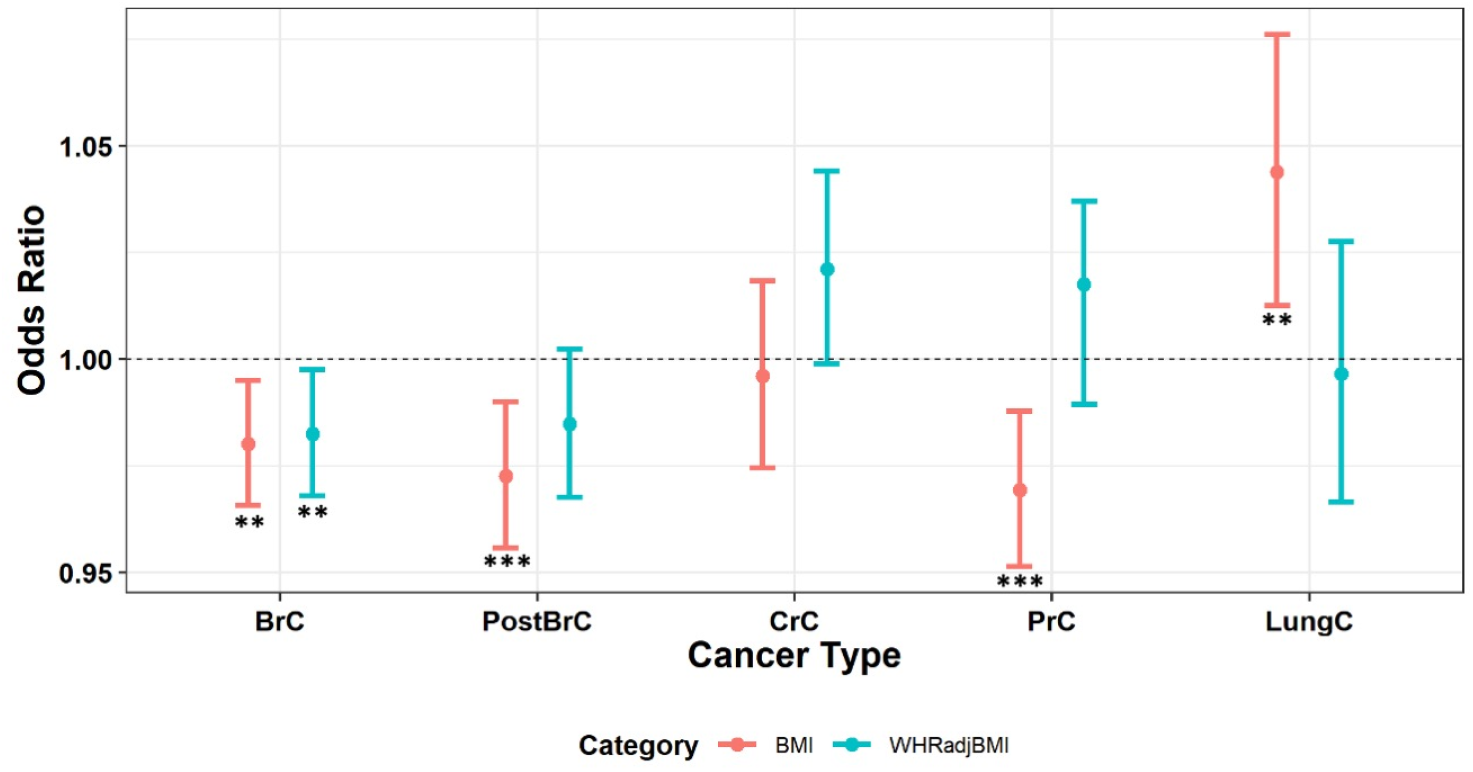
The association analysis results between PGS for BMI and WHRadjBMI and cancer in the UK Biobank. Legend: BrC=overall breast cancer, PostBrC=post-menopausal breast cancer, PrC=prostate cancer, CrC=colorectal cancer, LungC=lung cancer. Estimates which withstood Bonferroni corrections (*P*<0.05/10=0.005) are marked with triple asterisks (***), double asterisks (**) for 0.05 ≥ *P* > 0.005

**Table 1.** Association results between BMI and WHRadjBMI PGS and cancers in UK Biobank

| **Cancer** | **BMI PGS** |  | **WHRadjBMI PGS** |  |
| --- | --- | --- | --- | --- |
|  | **OR(95%CI)** | **P** | **OR(95%CI)** | **P** |
| BrC | 0.98(0.97-0.99) | 0.0086 | 0.98(0.97-0.99) | 0.021 |
| PostBrC | 0.97(0.96-0.99) | 0.00203 | 0.98(0.97-1.0023) | 0.089 |
| PrC | 0.97(0.95-0.99) | 0.0012 | 1.018(0.99-1.037) | 0.074 |
| CrC | 0.99(0.97-1.02) | 0.73 | 1.021(0.99-1.044) | 0.062 |
| LungC | 1.044(1.013-1.076) | 0.0057 | 0.99(0.97-1.027) | 0.82 |

Legend: BrC=breast cancer, PostBrC=post-menopausal breast cancer, PrC=prostate cancer, CrC=colorectal cancer, LungC=lung cancer, OR(95%CI)=odds ratio and the lower and upper confidence intervals

WHRadjBMI PGS (274 SNPs) was not significantly associated with any of the cancers. However, a nominal association was identified with overall breast cancer (OR[95%CI]=0.98[0.97-0.99], *P*=0.021). WHRadjBMI PGS had a trend towards positive association with prostate cancer (OR[95%CI]=1.018[0.99-1.037, *P*=0.074) and colorectal cancer (OR[95%CI]=1.021[0.999-1.044], *P*=0.062) (**Figure 4, Table 1**). The direct relationship detected through suggestive association between WHRadjBMI and prostate cancer is noteworthy as it is contrary to the BMI PGS results. Additionally, we calculated the associations between BMI PGS and cancer while stratifying the data by BMI categories. On average, the strength of associations for BMI PGS was higher for overweight and obese individuals (BMI higher than 25 kg/m2) (**Table S5**). We then used smoking status as a proxy for tobacco use in the lung cancer association tests, but we did not identify any significant association among current and previous smokers (**Table S6**). However, there was a nominal inverse association between WHRadjBMI PGS and lung cancer risk among individuals who had never smoked (OR[95%CI]=0.92[0.85-1.00], *P*=0.046).

### Causality using Mendelian randomization

In this study we investigated the bi-directional causal relationship between two adiposity phenotypes and three cancers. Our results demonstrate a significant protective causal effect of BMI on prostate cancer (OR_IVW_[95%CI]=0.993[0.988-0.998], *P*=4.19×10^−3^, 574 SNPs) (**Table 2**). Additionally, the sensitivity analysis done using the weighted median (W.Med) MR method agreed with the MR-IVW results (OR_W.Med_[95%CI]=0.993[0.985-0.9996], *P*=0.039, 574 SNPs). The scatter plot and odds ratio plots are shown in **Figure 5a**. The MR Egger intercept test suggested no evidence of pleiotropy among the selected SNPs in the BMI to prostate cancer test (Egger intercept =-46.47, *P*=0.713). The Cochran’s Q test indicated significant heterogeneity in this association (Q_IVW_=863.64, *P*=3.93×10^−14^) (**Table S7**). However, in the reverse direction from prostate cancer to BMI, there was no evidence of causality. All other BMI to cancer direction tests (and their reverse) did not show evidence of causality (**Figure S2 and S3, Table S7**).

**Table 2.** Mendelian randomization results with cancers as the exposure and adiposity measures as the outcome variable using the five different methods

| Exposure | Outcome | Inverse variance weighted |  | MR Egger |  | Weighted median |  | Simple mode |  | Weighted mode |  |
| --- | --- | --- | --- | --- | --- | --- | --- | --- | --- | --- | --- |
|  |  | OR (95% CI) | P | OR (95% CI) | P | OR (95% CI) | P | OR (95% CI) | P | OR (95% CI) | P |
| BMI | BrC | 1.000<br>(0.995-1.005) | 0.897 | 0.985<br>(0.971-1.000) | 0.051 | 0.996<br>(0.988-1.003) | 0.266 | 0.944<br>(0.968-1.020) | 0.634 | 0.994<br>(0.977-1.011) | 0.462 |
| BMI | PrC | **0.993**<br>**(0.988-0.998)** | **0.0042** | 0.995<br>(0.982-1.009) | 0.473 | **0.993**<br>**(0.985-0.999)** | **0.039** | 0.984<br>(0.960-1.009) | 0.22 | 0.995<br>(0.981-1.009) | 0.491 |
| BMI | CrC | 1.000<br>(0.998-1.002) | 0.92 | 1.001<br>(0.996-1.006) | 0.682 | 1.000<br>(0.997-1.003) | 1 | 0.999<br>(0.989-1.008) | 0.768 | 1.000<br>(0.995-1.005) | 0.923 |
| WHRadjBMI | BrC | **0.990**<br>**(0.983-0.997)** | **0.0068** | 1.000<br>(0.982-1.017) | 0.974 | 0.991<br>(0.981-1.002) | 0.105 | 1.003<br>(0.974-1.033) | 0.83 | 0.993<br>(0.978-1.008) | 0.338 |
| WHRadjBMI | PrC | 1.0046<br>(0.998-1.011) | 0.179 | **1.016**<br>**(1.00018-1.032)** | **0.048** | 1.007<br>(0.999-1.016) | 0.094 | 1.018<br>(0.990-1.045) | 0.209 | **1.022**<br>**(1.00067-1.038)** | **0.0053** |
| WHRadjBMI | CrC | 1.002<br>(0.994-1.004) | 0.125 | 1.000 (0.995-1.006) | 0.885 | 1.000<br>(0.996-1.004) | 0.917 | 0.995<br>(0.984-1.006) | 0.391 | 1.000<br>(0.994-1.006) | 0.988 |

Legend: BrC=breast cancer, PostBrC=post-menopausal breast cancer, PrC=prostate cancer, CrC=colorectal cancer, LungC=lung cancer, OR(95%CI)=odds ratio and the lower and upper 95% confidence intervals. Causal estimates with $P<0.05$ are shown in bold.

**Figure 5.**
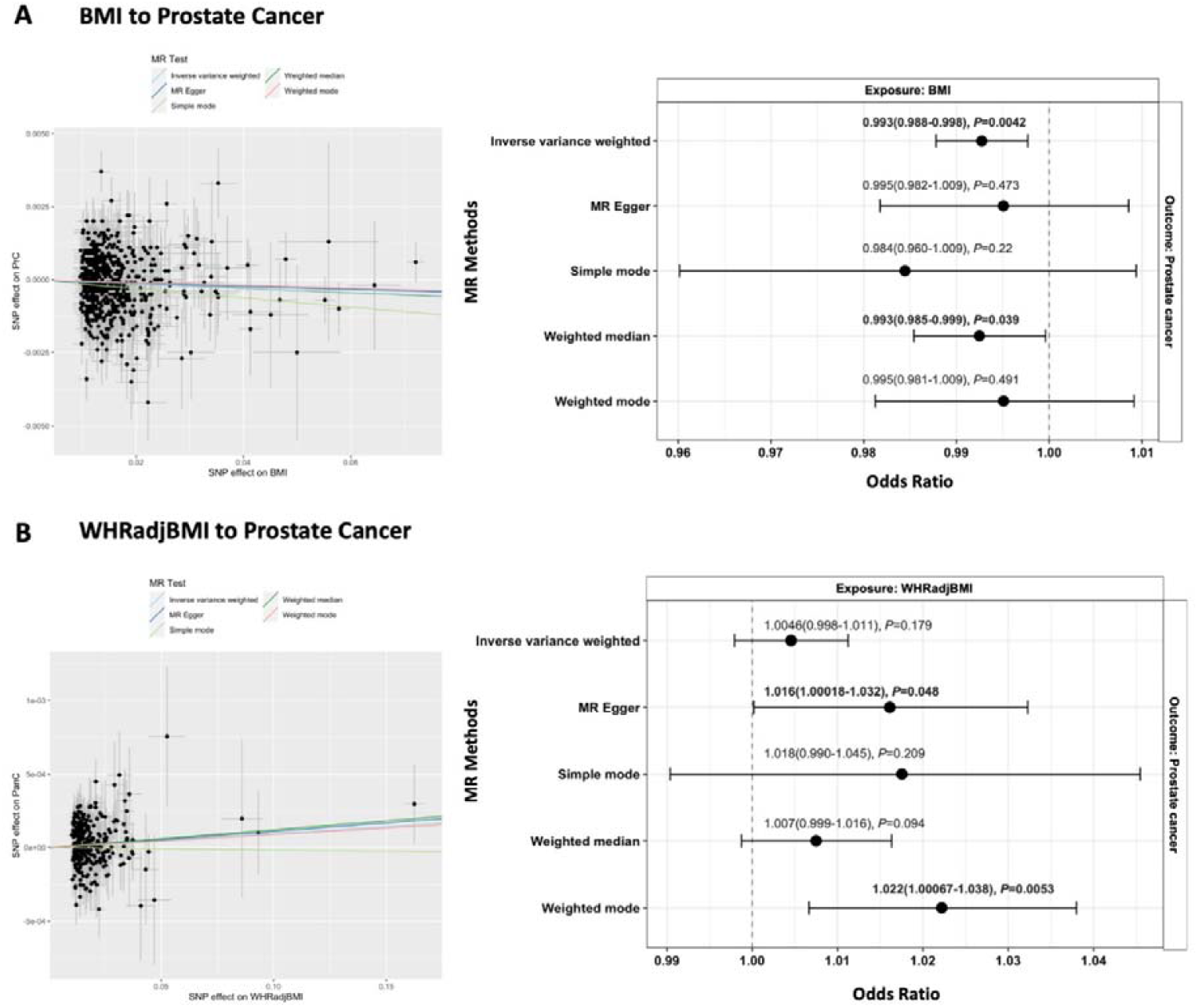
Scatter and forest plots from Mendelian Randomization analysis for A) BMI to prostate cancer MR, B) WHRadjBMI to prostate cancer. Legend: The scatter plots on the left also include the intercepts of the various MR methods used. The forest plots show the MR estimates for each of the methods used with the odds ratio, 95% confidence intervals and *P* values annotated. Causal estimates with *P*<0.05 are highlighted in bold.

In contrast, the WHRadjBMI to cancer causal test suggested a nominal causal risk effect of WHRadjBMI on prostate cancer risk (**Table 2**) based on the two MR tests used as sensitivity analyses (OR_MR-Egger_[95%CI]=1.016[1.00018-1.032], *P*=0.048, 284 SNPs; OR_W.Mode_[95%CI]=1.022[1.0067-1.038, *P*=0.0053, 284 SNPs (**Figure 5b, Table 2**). Additionally, there was a nominal causal protective effect of WHRadjBMI on overall breast cancer based on the MR-IVW test (OR_IVW_[95%CI]=0.99[0.98-0.997], *P*=0.0068, 284 SNPs) (**Table 2, Table S7**). However, the sensitivity tests were insignificant. None of the other WHRadjBMI to cancer direction tests (and their reverse) were significant (**Figure S4 and S5, Table S7**).

## DISCUSSION

In this study, we provide evidence that higher overall adiposity, measured with BMI, may confer men a protective advantage against prostate cancer, using genetic correlation analyses, polygenic scores, and Mendelian randomization. Conversely, higher abdominal adiposity, determined with WHRadjBMI, may be a risk factor for prostate cancer in men. We also report a nominal protective causal effect of central obesity on breast cancer. Additionally, we show nominal genetic correlation between higher abdominal adiposity and lung cancer. Still, among individuals with no history of tobacco use, higher abdominal adiposity appears to be protective of lung cancer. The association between higher BMI and lower prostate cancer risk is in line with observational studies(7,27). These results are not surprising since it is well-documented that type 2 diabetes (T2D) appears to be protective of prostate cancer(28,29), with obesity being a likely risk factor(30) for T2D. The mechanisms behind the inverse association between prostate cancer and high BMI are yet to be fully defined but several factors need to be considered when interpreting this finding. First, the impact of height on BMI must be put into perspective while highlighting the inverse association between high BMI and prostate cancer risk. BMI is defined by dividing weight by the square of height and as such, tall men may present with lower BMI despite having high body weight. Furthermore, height is an established risk factor for prostate cancer with elevated height linked to increased growth factors such as the insulin-like growth factor 1 (IGF-1)(31,32). Additionally, it has been suggested that the low serum testosterone levels typical among obese men(33) may be responsible as it has been shown that elevated free testosterone levels in men are associated with an increased risk of prostate cancer(34). Moreover, the association between higher BMI and prostate cancer may be influenced by tumour characteristics. For instance, higher BMI has been shown to increase the risk of aggressive prostate cancer and mortality(27,35,36), while higher BMI has been associated with reduced risk of overall prostate cancer and non-aggressive prostate cancer tumours(27,36).

Conversely, we also show a nominal positive association between higher abdominal adiposity and prostate cancer, in line with epidemiological evidence(9). Abdominal adiposity is associated with markers of metabolic dysfunction, such as insulin resistance and impaired glucose metabolism(3). These results therefore suggest that while total adiposity appears to be protective of prostate cancer, WHRadjBMI is a better predictor of prostate cancer risk, similar to epidemiological observations(37). However, the impact of body height once again warrants a consideration while interpreting this observation. By adjusting for BMI, the effect of height which is an established risk factor for prostate cancer(31) is excluded and could potentially explain the opposite results got for BMI and WHRadjBMI in respect to prostate cancer.

Furthermore, our results indicate that abdominal adiposity, independent of BMI, relates to cancer risk in an opposite manner for sex-specific cancers. Particularly, higher abdominal adiposity in men is a risk factor for prostate cancer, and in contrast, higher abdominal adiposity in women appears to offer a protective advantage against overall breast cancer. However, the latter is contrary to what is seen from observational studies(38). In this study, we considered cross-sectional adiposity measures, which do not reflect the obesity trajectories in women’s lifetime, a factor that might influence the exposure to diverse hormones. Other factors such as adiposity at age of menarche could have prolonged effects into adulthood and could in part explain the negative association between central adiposity and breast cancer. Indeed, higher BMI in early childhood and adolescence have been associated with decreased risk of breast cancer(39). Further work to unravel how early-life adiposity and sex hormones influence risk in an opposite direction for sex-specific cancers is needed.

In addition, the observation from genetic correlation and polygenic scores that higher BMI PGS directly relates to increased lung cancer risk is contrary to what is observed in epidemiological studies(4,40,41). Nevertheless, the significance of this positive association declined once we adjusted for smoking status as a proxy for tobacco use that is associated with lower BMI. Interestingly, this data shows that among individuals who reported to have no history of smoking, higher WHR independent of BMI was protective of lung cancer risk. Moreover, previous findings suggest that higher abdominal adiposity is a risk factor for lung cancer among current smokers(40,41). The mechanism behind this observation is unclear. However, it has been shown that preclinical lung cancer at baseline among current smokers may be associated with higher central adiposity(40).

Despite the lack of statistically significant findings in either PGS and MR analyses for colorectal cancer, there was a trend towards positive association between WHRadjBMI and colorectal cancer in PGS analyses whereas with BMI there was no evidence of association. This is consistent with epidemiological evidence suggesting that abdominal obesity is a stronger predictor for colorectal cancer than overall body weight(42,43). Insulin resistance and consequent hyperinsulinemia, and dyslipidemia associated with metabolic dysfunction have been describes as potential pathophysiology underlying the association with abdominal obesity and colorectal cancer risk(44,45).

This study has some limitations to be considered. Cancer subtypes based on tumour aggressiveness, or biomarker specificity are not considered based on the cancer definitions utilised in this study. A range of cancer subgroups would be characterised by different properties that could relate in different ways to adiposity measures. Future work on this analysis will work on partitioning adiposity GWAS variants into groups based on their apparent mechanistic functions. This would aide in explaining underlying co-morbidity between adiposity and cancer with specific biological pathways such as insulin resistance, inflammation, lipid metabolism, and immunity, in mind. Additionally, the low sample sizes for cancers in our analyses undermine the statistical power for cancer GWAS compared to that of adiposity phenotypes(10–15). Consequently, the precision of the risk estimates derived from Mendelian randomization results was affected and as such validation using larger datasets for cancer phenotypes is required. Finally, due to the lack of publicly available data for certain cancers such as lung and post-menopausal breast cancers, we did not assess causality between these cancers and adiposity.

Overall, we highlight the importance of assessing different adiposity measures in the context of cancer risk by employing analysis of two routine yet different measures of adiposity, BMI and WHRadjBMI, dissecting the relationships between them. We conclude that metabolic dysfunction, through WHRadjBMI, rather than just overall adiposity may be more informative of cancer risk for prostate cancer. However, the impact of height may play a role in the complex relationship between obesity and prostate cancer. Additionally, the differences in risk conferred by central adiposity may occur in opposite direction for sex-specific cancers as seen with prostate and breast cancer. Validation using larger cancer-focussed datasets would be required to confirm our findings.

## Data Availability

All data produced in the present study are available upon reasonable request to the authors

## ACKNOWLEDGEMENTS

We would like to thank the Genetic Investigation of Anthropometric Traits (GIANT) consortium for enabling access to their GWAS summary statistics which we have used in our PGS and MR analyses. We would also like to appreciate the participants and staff of the UK Biobank for their valuable contribution.

