## Supplementary for "Genetic relationships and causality between overall and central adiposity and breast, prostate, lung and colorectal cancer"

**Supplementary Materials**

**UK Biobank Genome-wide association studies**

We performed single phenotype GWAS in the UKBB phenotypes using the BOLT-LMM version 2.3 software^1^ which implements a linear mixed model (LMM) association testing. Consequently, as a result of applying a linear mixed model, related individuals in the UKBB were included in the association analyses. The standard BOLT-LMM v2.3 infinitesimal model was used. Among the 487,409 individuals with genetic data, the genetic data was filtered based on MAF > 0.01, imputation score > 0.4, Hardy-Weinberg Equilibrium (HWE) P-value >1x10^-6^ and per SNP variant missingness <0.015. As a result, 471,095 individuals passed these filters. We included age, sex, genotyping array and six principal components (PCs) as covariates in the LMM for BMI, WHRadjBMI, colorectal cancer (CrC) and lung cancer (LungC). For the sex-specific cancer phenotypes breast (BrC), post-menopausal breast (PostBrC) and prostate (PrC) cancers, sex was not included as a covariate. Moreover, BMI was included as an additional covariate in WHR association testing to obtain the WHRadjBMI phenotype. The threshold for statistically significant genome-wide signals (SNPs) was 5x10^-8^. Manhattan plots for the association results are show in **Supplementary Figures 6-9**.

**Supplementary Figures**

**Supplementary Figure 1**. Summary of Mendelian Randomization tests performed, including the number of genetic instruments (SNPs) available from published GWAS for each test

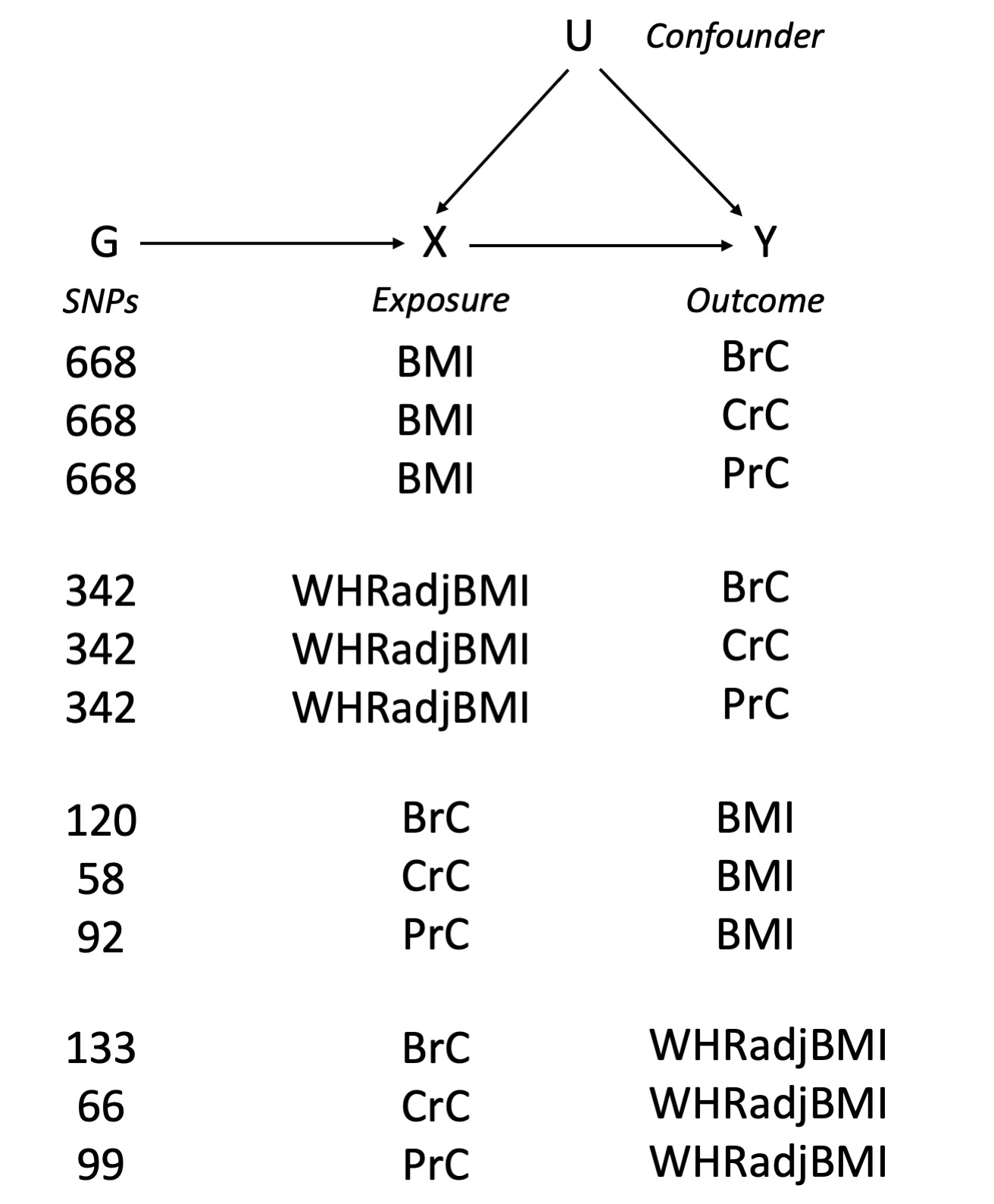

**Supplementary Figure 2**. Scatter and forest plots for the BMI to cancer direction tests

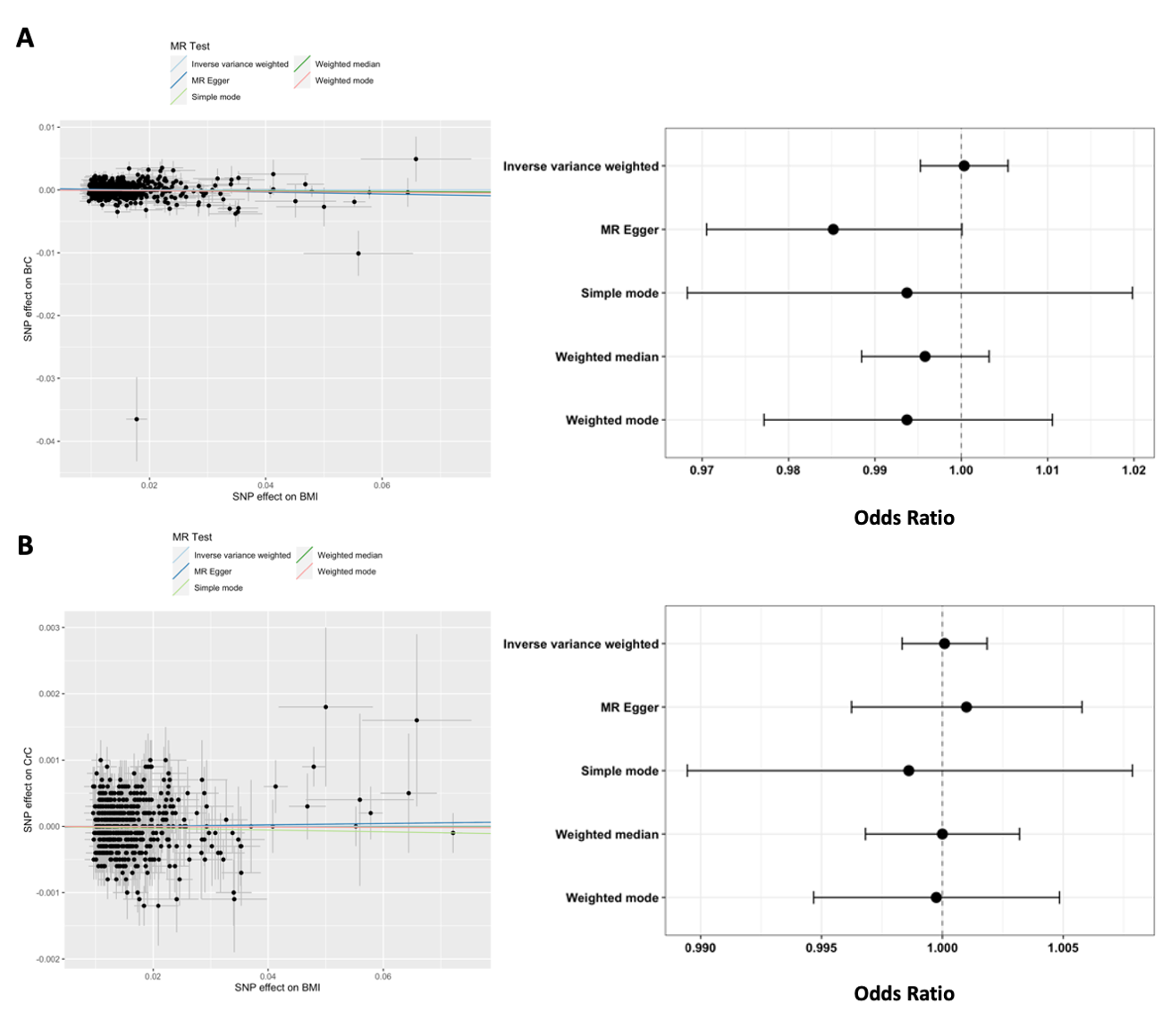

Legend: A=BMI to Breast cancer, B=BMI to colorectal cancer

**Supplementary Figure 3**. Scatter and forest plots for the cancer to BMI direction tests

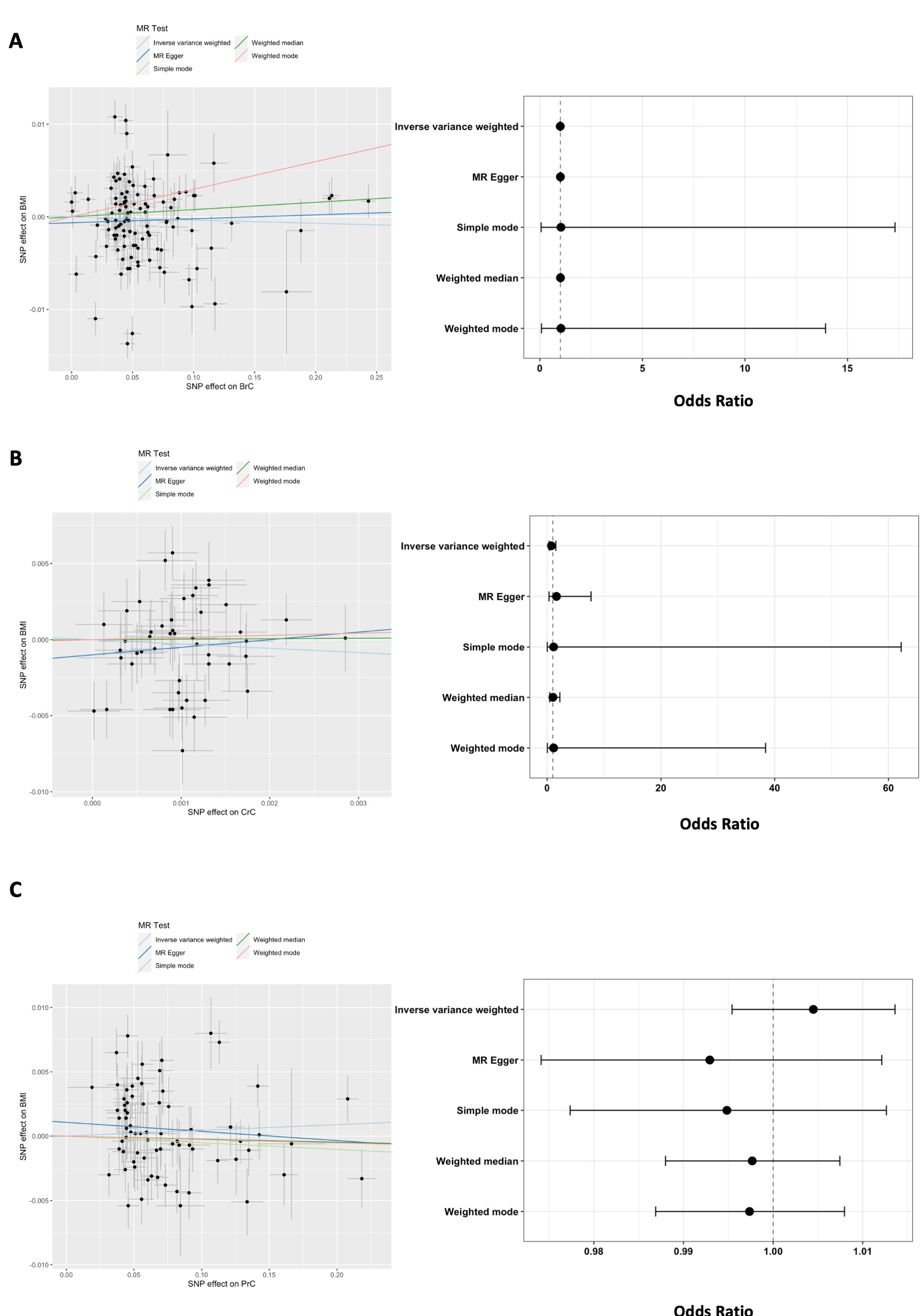

Legend: A= Breast cancer to BMI, B=Colorectal cancer to BMI, C=Prostate cancer to BMI

**Supplementary Figure 4**. Scatter and forest plots for the WHRadjBMI to cancer direction tests

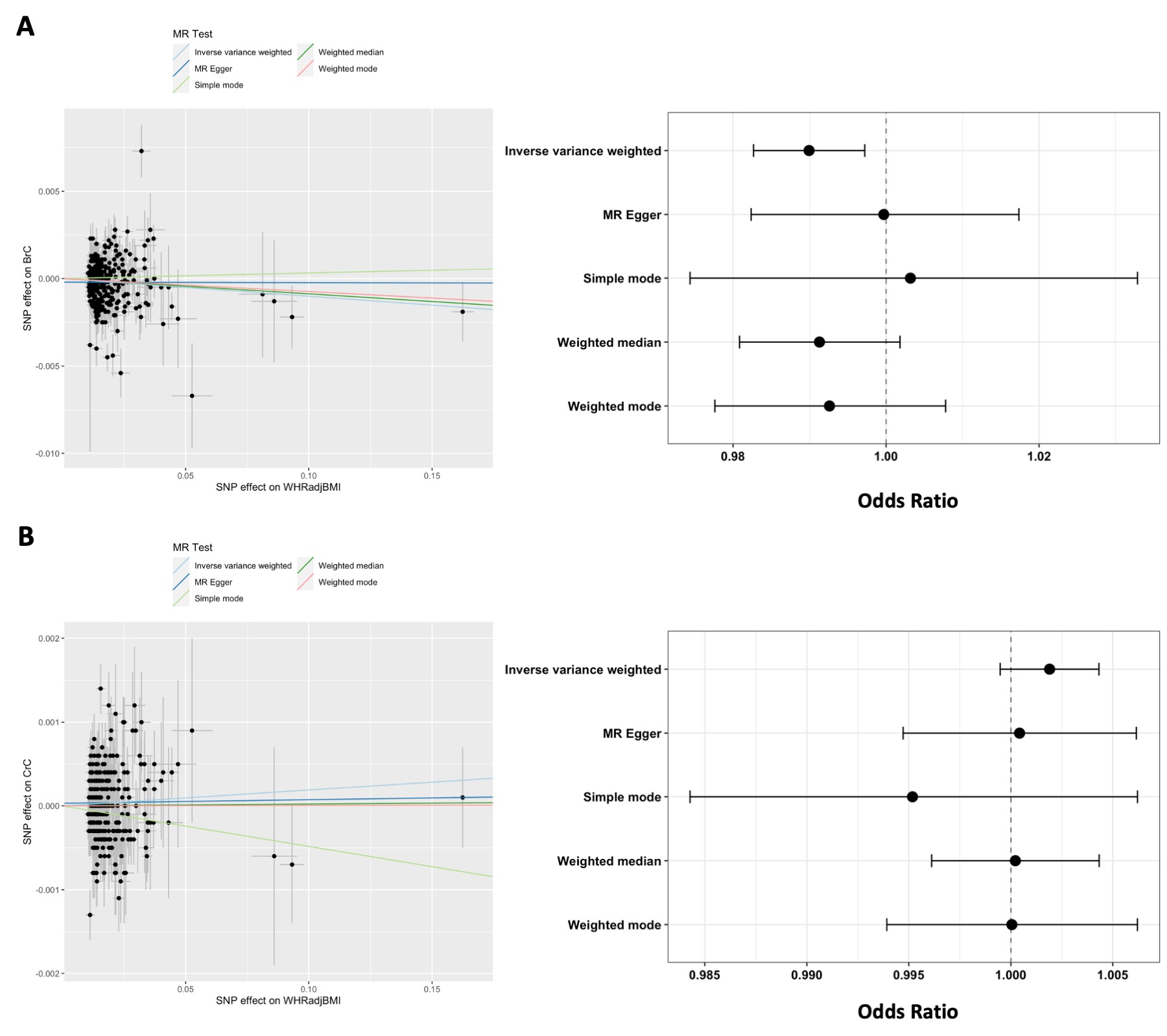

Legend: A=WHRadjBMI to Breast cancer, B=WHRadjBMI to Colorectal cancer

**Supplementary Figure 5**. Scatter and forest plots for the cancer to WHRadjBMI direction tests

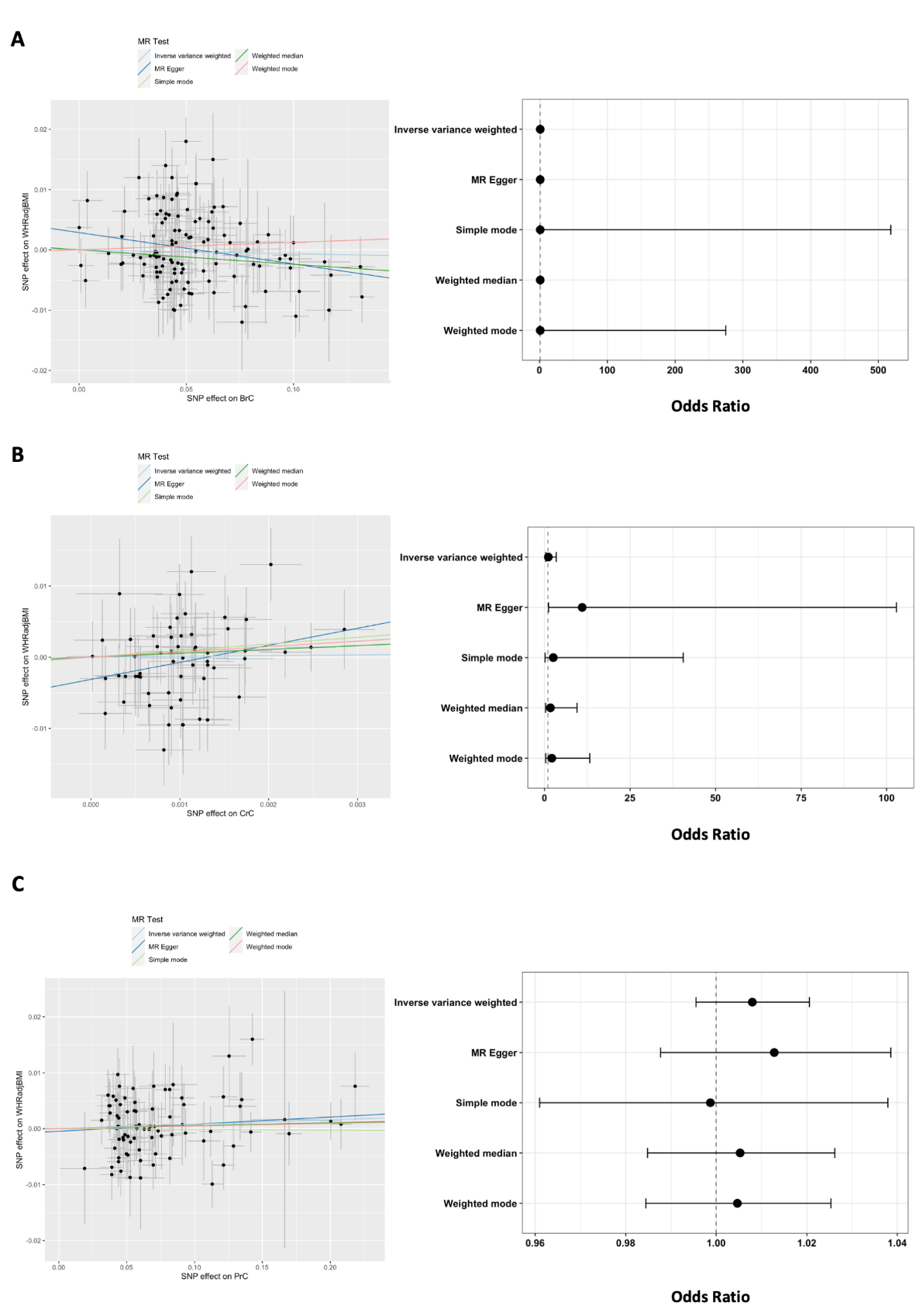

Legend: A= Breast cancer to WHRadjBMI, B=Colorectal cancer to WHRadjBMI, C=Prostate cancer to WHRadjBMI

**Supplementary figure 6**. Manhattan plots of overall breast cancer (top) and post-menopausal breast cancer (bottom) GWAS in UK Biobank. The red horizontal line shows genome-wide significance threshold (P<5x10^-8^). The dashed grey line shows suggestive significance threshold (P<1x10^-5^)

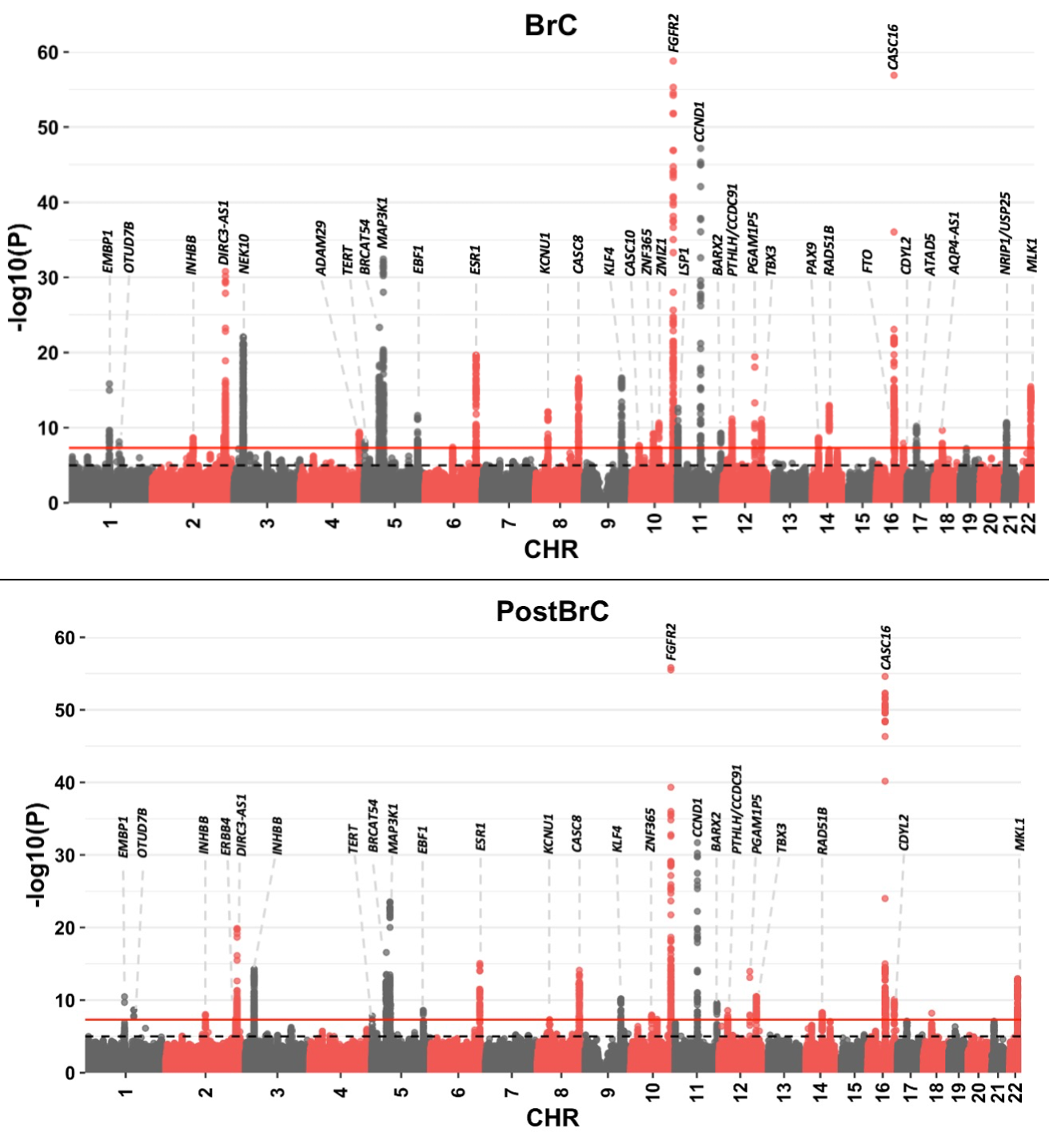

**Supplementary figure 7**. Manhattan plots of prostate cancer (top) and colorectal cancer (bottom) GWAS in UK Biobank. The red horizontal line shows genome-wide significance threshold (P<5x10^-8^). The dashed grey line shows suggestive significance threshold (P<1x10^-5^)

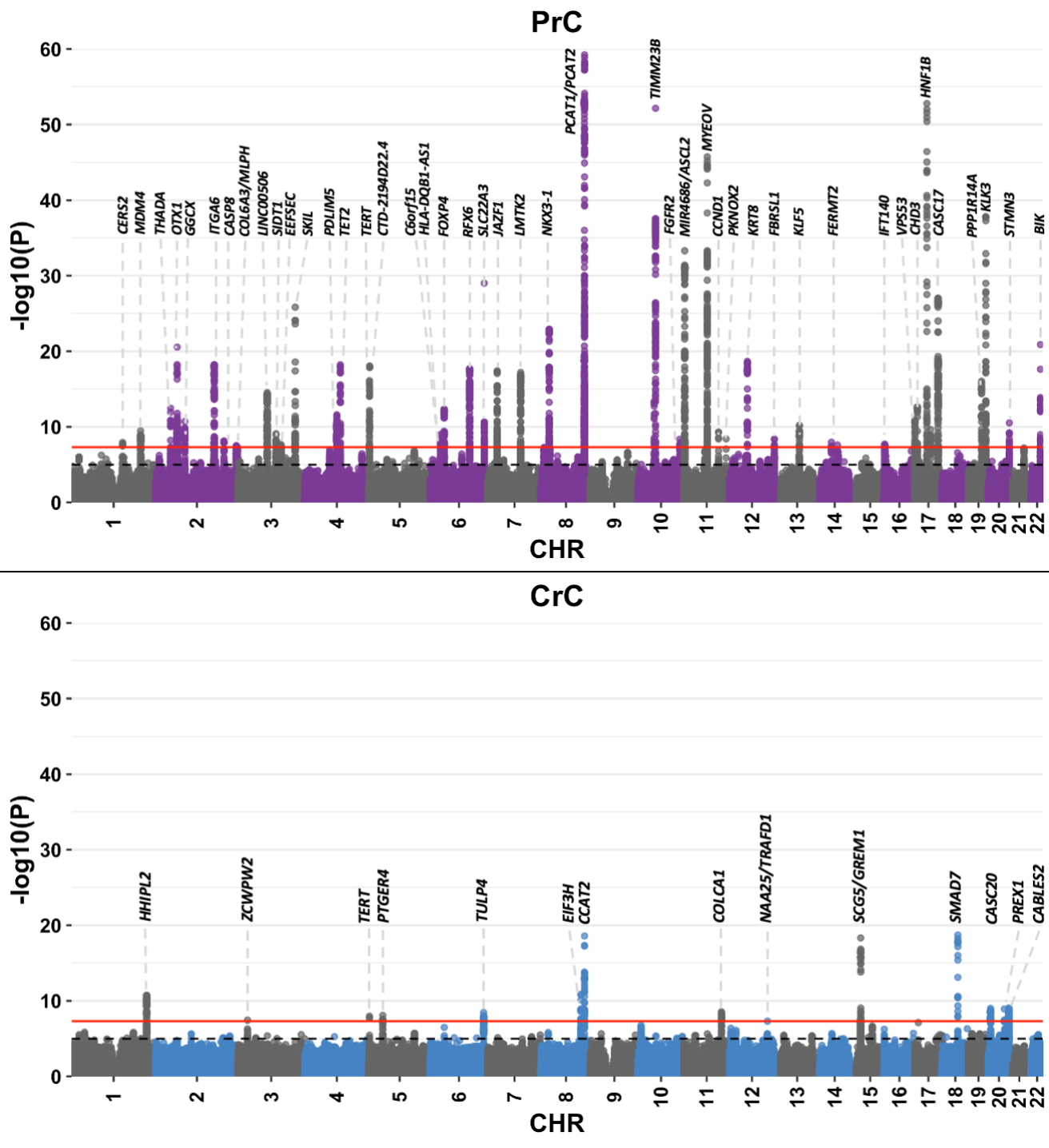

**Supplementary figure 8**. Manhattan plot of lung cancer GWAS in UK Biobank. The red horizontal line shows genome-wide significance threshold (P<5x10^-8^). The dashed grey line shows suggestive significance threshold (P<1x10^-5^)

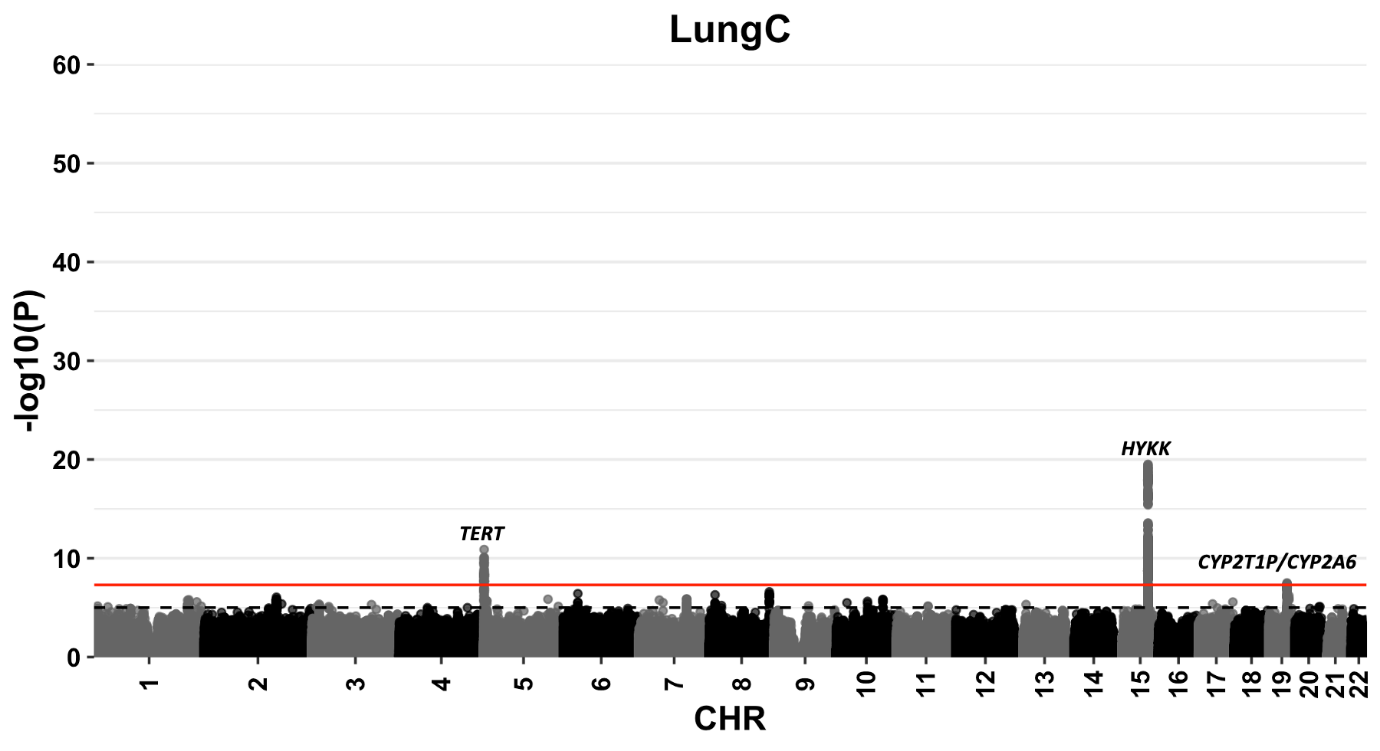

**Supplementary figure 9**. Manhattan plots of BMI (top) and WHRadjBMI cancer (bottom) GWAS in UK Biobank. The red horizontal line shows genome-wide significance threshold (P<5x10^-8^).

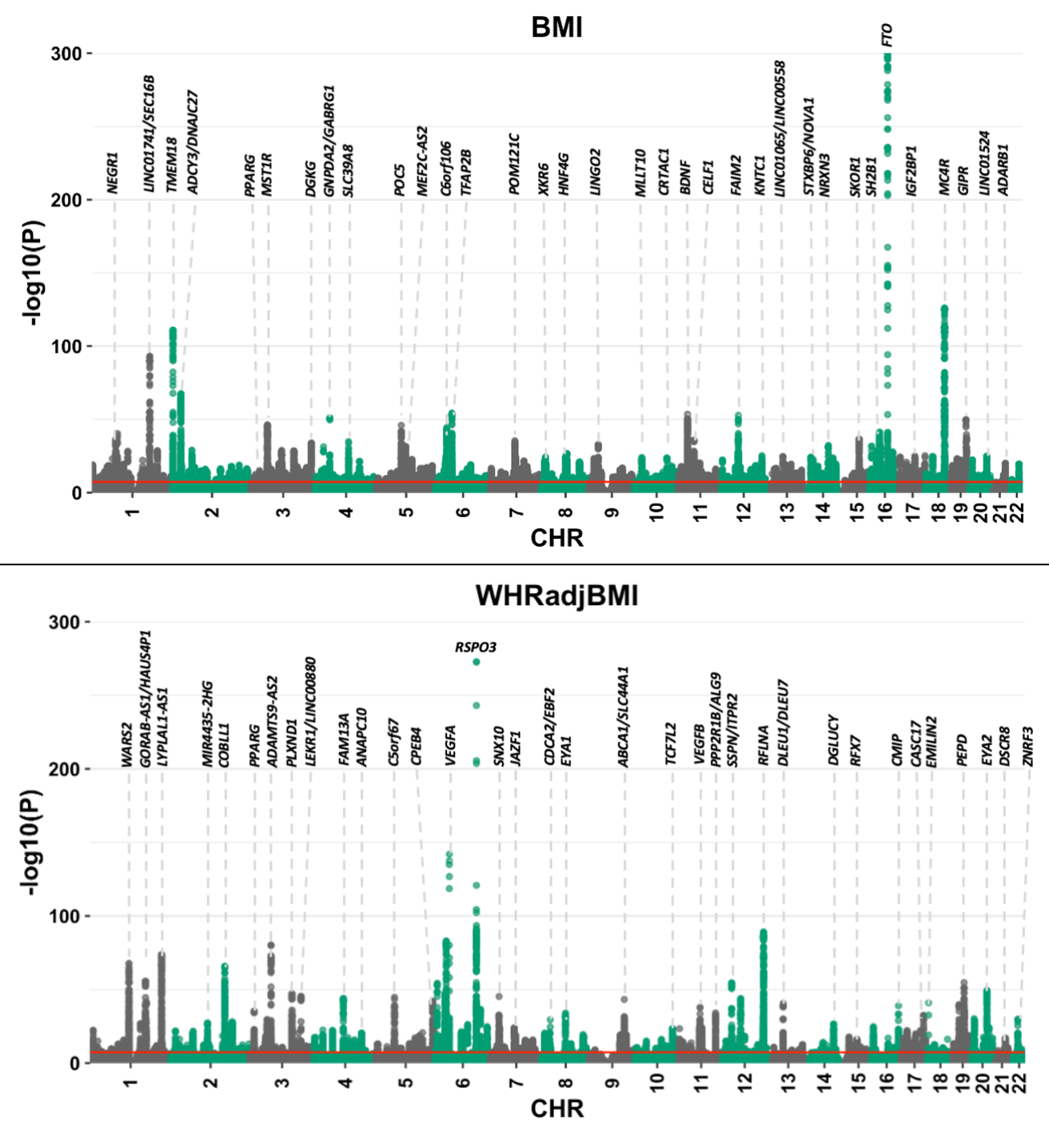

**Supplementary Tables**

**Supplementary table 1. Phenotype definition criteria for cancer phenotypes in UK Biobank**

| **Cancer** | **UK Biobank field description** | **ICD-10 Codes** |
| --- | --- | --- |
| Overall breast cancer (BrC) | Have been diagnosed with breast cancer AND Breast cancer is the first cancer diagnosed OR Death cause is breast cancer | C50 |
| Post-menopausal breast cancer (Post-BrC) | Have been diagnosed with breast cancer AND Breast cancer is the first cancer diagnosed OR Death cause is breast cancer AND Self-reported menopause status | C50 and X2724 (Menopause status) |
| Colorectal cancer (CrC) | Have been diagnosed with colon and rectal cancer AND colon and rectal cancers are the first cancers diagnosed OR Death cause is colon and rectal cancers | C18-C21 |
| Prostate cancer (PrC) | Have been diagnosed with prostate cancer AND prostate cancer is the first cancer diagnosed OR Death cause is prostate cancer | C61 |
| Lung cancer (LungC) | Have been diagnosed with lung cancer AND lung cancer is the first cancer diagnosed OR Death cause is lung cancer | C34 |

**Supplementary table 2. UK Biobank study participant numbers for obesity and cancer phenotypes**

| **Phenotype** | **Cases** | **Controls** | **Total** |
| --- | --- | --- | --- |
| BrC | 18,676 | 230,571 | 249,247 |
| Post-BrC | 13,355 | 235,804 | 249,159 |
| PrC | 11,825 | 197,912 | 209,737 |
| CrC | 8,201 | 450,693 | 458,894 |
| LungC | 4,237 | 455,010 | 459,247 |
| BMI | NA | NA | 457,270 |
| WHRadjBMI | NA | NA | 457,270 |

**Supplementary table 3. Genetic correlation estimates between BMI/WHRadjBMI and cancer in UK Biobank**

| **Cancer** | **Adiposity trait** | **Genetic correlation (rG)** | **SE** | **P** |
| --- | --- | --- | --- | --- |
| BrC | BMI | -0.035 | 0.03 | 0.236 |
| Post-BrC | BMI | -0.0803 | 0.03 | 0.014 |
| PrC | BMI | -0.076 | 0.028 | 0.0075 |
| CrC | BMI | 0.0089 | 0.039 | 0.82 |
| LungC | BMI | 0.18 | 0.056 | 0.0014 |
| BrC | WHRadjBMI | 0.009 | 0.028 | 0.75 |
| Post-BrC | WHRadjBMI | -0.017 | 0.035 | 0.63 |
| PrC | WHRadjBMI | 0.025 | 0.03 | 0.408 |
| CrC | WHRadjBMI | 0.103 | 0.043 | 0.017 |
| LungC | WHRadjBMI | 0.159 | 0.058 | 0.0065 |

**Supplementary table 4. SNP heritability estimates of BMI/WHRadjBMI and cancer in UK Biobank**

| **Phenotype** | **h2** | **SE** |
| --- | --- | --- |
| BrC | 0.0323 | 0.0041 |
| Post-BrC | 0.0215 | 0.003 |
| PrC | 0.0441 | 0.005 |
| CrC | 0.0072 | 0.0013 |
| LungC | 0.0036 | 0.0011 |
| BMI | 0.2459 | 0.0072 |
| WHRadjBMI | 0.1343 | 0.0066 |

**Supplementary table 5. BMI PRS association with UK Biobank cancer by BMI categories**

|  | **BMI Class** | **Case/Control** | **OR (95%CI)** | **P** |
| --- | --- | --- | --- | --- |
| BrC | Underweight | 83/1775 | 0.94 (0.78-1.13) | 0.497 |
|  | Normal | 4,796/93,114 | 0.98 (0.96-1.01) | 0.178 |
|  | Pre-obesity | 5,277/86,114 | 0.98 (0.95-1.00) | 0.053 |
|  | **Obesity** | **3,164/54,072** | **0.96 (0.93-0.99)** | **0.012** |
| Post-BrC | Underweight | 124,1,734 | 0.87 (0.70-1.09) | 0.242 |
|  | Normal | 6,872/91,058 | 0.97 (0.94-1.00) | 0.029 |
|  | Pre-obesity | 7,188/84,240 | 0.98 (0.95-1.01) | 0.161 |
|  | **Obesity** | **4,436/52,830** | **0.96 (0.92-0.99)** | **0.017** |
| PrC | Underweight | 18.456 | 0.90 (0.55-1.47) | 0.68 |
|  | Normal | 2,926/48,880 | 0.98 (0.94-1.02) | 0.351 |
|  | **Pre-obesity** | **6,128/97,260** | **0.98 (0.95-1.00)** | **0.066** |
|  | Obesity | 2,728/50,611 | 0.98 (0.94-1.02) | 0.412 |
| CrC | Underweight | 27/2,305 | 0.89 (0.60-1.32) | 0.569 |
|  | Normal | 2,440/147,476 | 0.98 (0.94-1.02) | 0.35 |
|  | Pre-obesity | 3,711/191,068 | 0.99 (0.96-1.03) | 0.65 |
|  | Obesity | 2,193/108,380 | 0.97 (0.93-1.02) | 0.215 |
| LungC | Underweight | 46/2,289 | 1.13 (0.84-1.50) | 0.417 |
|  | Normal | 1,293/148,528 | 1.04 (0.99-1.10) | 0.136 |
|  | **Pre-obesity** | **1,763/193,131** | **1.08 (1.03-1.14)** | **0.001** |
|  | Obesity | 1,105/109,592 | 1.01 (0.95-1.07) | 0.856 |

Associations with *p*<0.05 are shown in bold

**Supplementary table 6. Adiposity PRS association with lung cancer by smoking status as a proxy for tobacco use**

|  |  | **BMI PRS** | | **WHRadjBMI PRS** | |
| --- | --- | --- | --- | --- | --- |
| **Smoking status** | **Case/Control/N** | **OR (95%CI)** | **P** | **OR (95%CI)** | **P** |
| Previous smokers | 1,970/160,891 (162,861) | 1.02 (0.98-1.07) | 0.294 | 0.99 (0.95-1.04) | 0.737 |
| Current smokers | 1,652/46,242 (47,894) | 1.01 (0.96-1.06) | 0.839 | 1.03 (0.98-1.08) | 0.281 |
| Never smoked | 579/246,273  (246,852) | 1.01 (0.93-1.10) | 0.759 | 0.92 (0.85-1.00) | **0.046** |
| Previous + current smokers | 3,622/207,133 (210,755) | 1.02 (0.99-1.05) | 0.231 | 1.01 (0.98-1.04) | 0.638 |

Associations with *p*<0.05 are shown in bold

**Supplementary table 7. Detailed results of the Mendelian randomization analyses between adiposity and cancer phenotypes**

|  | | | **Inverse variance weighted** | | **MR Egger** | | **Weighted median** | | **Simple mode** | | **Weighted mode** | | **Heterogeneity** | **MR-Egger Intercept** | |
| --- | --- | --- | --- | --- | --- | --- | --- | --- | --- | --- | --- | --- | --- | --- | --- |
| **Exposure** | **Outcome** | **NSNPs** | **OR**  **(95% CI)** | **P** | **OR**  **(95% CI)** | **P** | **OR**  **(95% CI)** | **P** | **OR**  **(95% CI)** | **P** | **OR**  **(95% CI)** | **P** | **Q stat (P)** | **Intercept(SE)** | **P** |
| BMI | BrC | 576 | 1.000 (0.995-1.005) | 0.897 | 0.985 (0.971-1.000) | 0.051 | 0.996 (0.988-1.003) | 0.266 | 0.944 (0.968-1.020) | 0.634 | 0.994 (0.977-1.011) | 0.462 | 755.9 (5.34E-07) | 0.0003 (0.0001) | 0.034 |
| BMI | PrC | 574 | 0.993 (0.988-0.998) | 0.0042 | 0.995 (0.982-1.009) | 0.473 | 0.993 (0.985-0.999) | 0.039 | 0.984 (0.960-1.009) | 0.22 | 0.995 (0.981-1.009) | 0.491 | 863.64 (3.93E-14) | -45.47 (0.0001) | 0.713 |
| BMI | CrC | 575 | 1.000 (0.998-1.002) | 0.92 | 1.001 (0.996-1.006) | 0.682 | 1.000 (0.997-1.003) | 1 | 0.999 (0.989-1.008) | 0.768 | 1.000 (0.995-1.005) | 0.923 | 656.59 (0.0094) | -20.71 (0.00004) | 0.689 |
| BrC | BMI | 109 | 0.997 (0.985-1.008) | 0.557 | 1.004 (0.984-1.025) | 0.686 | 1.008 (0.998-1.017) | 0.1 | 1.030 (0.061-17.321) | 0.983 | 1.030 (0.076-13.925) | 0.982 | 510.98 (1.27E-53) | -0.006 (0.0007) | 0.372 |
| PrC | BMI | 74 | 1.004 (0.995-1.014) | 0.333 | 0.993 (0.974-1.102) | 0.471 | 0.998 (0.988-1.007) | 0.639 | 0.995 (0977-1.013) | 0.57 | 0.997 (0.987-1.008) | 0.626 | 194.16 (6.04E-13) | 0.001 1 (0.0008) | 0.184 |
| CrC | BMI | 48 | 0.752 (0.368-1.538) | 0.435 | 1.648 (0.352-7.729) | 0.529 | 1.031 (0.473-2.246) | 0.939 | 1.153 (0.021-62.240) | 0.944 | 1.153 (0.035-38.392) | 0.937 | 102.89 (4.71E-06) | -0.001 (0.0009) | 0.268 |
| WHRadjBMI | BrC | 284 | 0.990 (0.983-0.997) | 0.0068 | 1.000 (0.982-1.017) | 0.974 | 0.991 (0.981-1.002) | 0.105 | 1.003 (0.974-1.033) | 0.83 | 0.993 (0.978-1.008) | 0.338 | 529.41 (3.63E-17) | -0.0002 (0.0002) | 0.226 |
| WHRadjBMI | PrC | 284 | 1.0046 (0.998-1.011) | 0.179 | 1.016 (1.00018-1.032) | 0.048 | 1.007 (0.999-1.016) | 0.094 | 1.018 (0.990-1.045) | 0.209 | 1.022 (1.00067-1.038) | 0.0053 | 493.63 (1.32E-13) | -0.0002 (0.0002) | 0.119 |
| WHRadjBMI | CrC | 284 | 1.002 (0.994-1.004) | 0.125 | 1.000 (0.995-1.006) | 0.885 | 1.000 (0.996-1.004) | 0.917 | 0.995 (0.984-1.006) | 0.391 | 1.000 (0.994-1.006) | 0.988 | 410.68 (1.02E-06) | 0.00003 (0.00006) | 0.578 |
| BrC | WHRadjBMI | 117 | 0.993 (0.975-1.012) | 0.495 | 0.949 (0.910-0.991) | 0.018 | 0.976 (0.953-1.001) | 0.056 | 1.013 (0.002-518.382) | 0.997 | 1.013 (0.004-274.794) | 0.997 | 180.81 (1.11E-04) | 0.003 (0.001) | 0.022 |
| PrC | WHRadjBMI | 81 | 1.008 (0.996-1.021) | 0.211 | 1.013 (0.988-1.039) | 0.324 | 1.005 (0.985-1.026) | 0.616 | 0.999 (0.961-1.038) | 0.947 | 1.005 (0.984-1.025) | 0.655 | 79.36 (0.499) | -0.0005 (0.001) | 0.668 |
| CrC | WHRadjBMI | 59 | 1.113 (0.362-3.423) | 0.852 | 11.000 (1.176-102.88) | 0.04 | 1.728 (0.314-9.509) | 0.53 | 2.546 (0.016-40.531) | 0.511 | 2.126 (0.342-13.222) | 0.422 | 68.69 (0.159) | -0.003 (0.001) | 0.025 |
