## Supplementary for "Genetic relationships and causality between overall and central adiposity and breast, prostate, lung and colorectal cancer"

**STrengthening the REporting of Genetic Association studies (STREGA) reporting recommendations, extended from STROBE Statement**

| **Item** | **Item no** | **STROBE Guideline** | **Extension for Genetic Association Studies (STREGA)** | **Page no** |
| --- | --- | --- | --- | --- |
| **Title and Abstract** | 1 | (a) Indicate the study’s design with a commonly used term in the title or the abstract. |  | In the title on page 1, we mention highlight that the genetic relationships and causality between obesity and cancer are investigated in this work |
|  |  | (b) Provide in the abstract an informative and balanced summary of what was done and what was found. |  | The structured abstract on page 4 summarises in 200 words the study objectives, methods, results and conclusions of this study |
| **Introduction** |  |  |  |  |
| *Background rationale* | 2 | Explain the scientific background and rationale for the investigation being reported. |  | On page 6 and the first paragraph of page 7, we introduce the research problem and give evidence from published work. |
| *Objectives* | 3 | State specific objectives, including any pre-specified hypotheses | ***State if the study is the first report of a genetic association, a replication effort, or both.*** | The last paragraph of page 7 clearly states the objectives of the study. |
| **Methods** |  |  |  |  |
| *Study design* | 4 | Present key elements of study design early in the paper. |  |  |
| *Setting* | 5 | Describe the setting, locations and relevant dates, including periods of recruitment, exposure, follow-up and data collection. |  | The overview of the main study dataset is given on page 7 at the start of methods |
| *Participants* | 6 | (a) (a) **Cohort study –** Give the eligibility criteria, and the sources and methods of selection of participants. Describe methods of follow-up.   **Case–control study –** Give the eligibility criteria, and the sources and methods of case ascertainment and control selection. Give the rationale for the choice of cases and controls.  **Cross-sectional study –** Give the eligibility criteria, and the sources and methods of selection of participants. | ***Give information on the criteria and methods for selection of subsets of participants from a larger study, when relevant.*** | a) The UK Biobank resource is introduced in page 7. The inclusion criteria for cases and controls is summarised in supplementary table 1 |
|  |  | **(b) Cohort study** – For matched studies, give matching criteria and number of exposed and unexposed.  **Case–control study –** For matched studies, give matching criteria and the number of controls per case. |  |  |
| *Variables* | 7 | (a) Clearly define all outcomes, exposures, predictors, potential confounders, and effect modifiers. Give diagnostic criteria, if applicable. | ***(b) Clearly define genetic exposures (genetic variants) using a widely –used nomenclature system. Identify variables likely to be associated with population stratification (confounding by ethnic origin).*** | In polygenic scores analyses and Mendelian randomization, we include the exact number of variants for obesity and cancer phenotypes used (Pages 8, 9). |
| *Data sources measurement* | 8* | (a) For each variable of interest, give sources of data and details of methods of assessment (measurement). Describe comparability of assessment methods if there is more than one group. | ***(b) Describe laboratory methods, including source and storage of DNA, genotyping methods and platforms (including the allele calling algorithm used, and its version), error rates and call rates. State the laboratory /centre where genotyping was done. Describe comparability of laboratory methods if there is more than one group. Specify whether genotypes were assigned using all of the data from the study simultaneously or in smaller batches.*** | A flowchart summarising how the genetic variants used in polygenic scores analyses were derived is shown in Figure 1.  The source of genetic variants used as instrument variables in Mendelian randomization is described in page 9 |
| *Bias* | 9 | (a) Describe any efforts to address potential sources of bias. | ***(b) For quantitative outcome variables, specify if any investigation of potential bias resulting from pharmacotherapy was undertaken. If relevant, describe the nature and magnitude of the potential bias, and explain what approach was used to deal with this.*** | To account for potential sample overlap in polygenic scores analyses, we show how we exclude the effect of UK Biobank from obesity summary statistics. This is described in page 8 and also in Figure 1. |
| *Study size* | 10 | Explain how the study size was arrived at. |  | The study sample sizes are described in page 7 and supplementary table 2 |
| *Quantitative variables* | 11 | Explain how quantitative variables were handled in the analyses. If applicable, describe which groupings were chosen, and why. | ***If applicable, describe how effects of treatment were dealt with.*** |  |
| *Statistical methods* | 12 | (a) Describe all statistical methods, including those used to control for confounding. | ***State software version used and options (or settings) chosen.*** | 1. GWAS in UK Biobank was done using the *BOLT-LMM* v2.3 software as described in supplementary. QC done is described in supplementary. 2. Genetic correlation and heritability estimates were computed the LD Score (LDSC) regression tool. The QC for this method is described in page 7 in methods. 3. We constructed the obesity polygenic scores in *Plink* v1.90b4.1 and is mentioned on page 8 in the methods section. 4. Mendelian randomization analyses were done using the *Two Sample MR* R package version 0.5.6 and it is mentioned on page 9 in methods |
|  |  | (b) Describe any methods used to examine subgroups and interactions. |  | b) We describe in page 8 of methods how we perform polygenic scores analyses in UK Biobank by BMI categories. |
|  |  | (c) Explain how missing data were addressed. |  |  |
|  |  | (d) **Cohort study –** If applicable, explain how loss to follow-up was addressed.  **Case–control study –** If applicable, explain how matching of cases and controls was addressed.  **Cross-sectional study –** If applicable, describe analytical methods taking account of sampling strategy. |  |  |
|  |  | (e) Describe any sensitivity analyses. |  |  |
|  |  |  | ***(f) State whether Hardy- Weinberg equilibrium was considered and, if so, how.*** | f) For UK Biobank GWAS, we used HWE p-value threshold of p>1x10-6 in *BOLT-LMM* software as described in supplementary methods. |
|  |  |  | ***(g) Describe any methods used for inferring genotypes or haplotypes.*** |  |
|  |  |  | ***(h) Describe any methods used to assess or address population stratification.*** | h) In UK Biobank GWAS, and polygenic scores analyses we adjust for principal components (PCs) as described in supplementary methods and main methods |
|  |  |  | ***(i) Describe any methods used to address multiple comparisons or to control risk of false positive findings.*** | i) We used the genome-wide significance threshold (5x10-8) to define significant signals in GWAS as described in supplementary methods  For polygenic scores and Mendelian randomization, we include Bonferroni adjusted p-values in main methods section pages 8,9 |
|  |  |  | ***(j) Describe any methods used to address and correct for relatedness among subjects.*** | j) In UK Biobank GWAS, we used the BOLT-LMM software which considers any sample relatedness as mentioned in supplementary methods. |
| **Results** |  |  |  |  |
| *Participants* | 13* | (a) Report the numbers of individuals at each stage of the study – e.g. numbers potentially eligible, examined for eligibility, confirmed eligible, included in the study, completing follow-up and analysed. | ***Report numbers of individuals in whom genotyping was attempted and numbers of individuals in whom genotyping was successful.*** | a) We outline the case-control numbers for each cancer defined in UK Biobank in Supplementary Table 2 |
|  |  | (b) Give reasons for non-participation at each stage. |  |  |
|  |  | (c) Consider use of a flow diagram. |  |  |
| *Descriptive data* | 14* | (a) Give characteristics of study participants (e.g. demographic, clinical, social) and information on exposures and potential confounders. | ***Consider giving information by genotype.*** |  |
|  |  | (b) Indicate the number of participants with missing data for each variable of interest. |  |  |
|  |  | (c) **Cohort study –** Summarize follow-up time, e.g. average and total amount. |  |  |
| *Outcome data* | 15* | **Cohort study –** Report numbers of outcome events or summary measures over time. | ***Report outcomes***  ***(phenotypes) for each genotype category over time*** |  |
|  |  | **Case–control study –** Report numbers in each exposure category, or summary measures of exposure. | ***Report numbers in each genotype category*** |  |
|  |  | **Cross-sectional study –** Report numbers of outcome events or summary measures. | ***Report outcomes (phenotypes) for each genotype category*** |  |
| *Main results* | 16 | (a) Give unadjusted estimates and, if applicable, confounder-adjusted estimates and their precision (e.g. 95% confidence intervals). Make clear which confounders were adjusted for and why they were included. |  | a) The results of UK Biobank GWAS are shown in supplementary data in form of Manhattan plots  Genetic correlation and heritability estimates in UK Biobank are describes in results section (page 10) of the main text and shown in supplementary tables 3 and 4 and in Figure 3.  Polygenic scores results are defined in the results section of the main text (Page 11) and in table 1 of the main text, supplementary tables 5 and 6. And Figure 4  Mendelian randomization results are described in the results section of the main text (page 12) and shown in table 1 of main text and supplementary table 7 as well as figure 5 |
|  |  | (b) Report category boundaries when continuous variables were categorized. |  |  |
|  |  | (c) If relevant, consider translating estimates of relative risk into absolute risk for a meaningful time period. |  |  |
|  |  |  | ***(d) Report results of any adjustments for multiple comparisons.*** |  |
| *Other analyses* | 17 | (a) Report other analyses done – e.g. analyses of subgroups and interactions, and sensitivity analyses. |  |  |
|  |  |  | ***(b) If numerous genetic exposures (genetic variants) were examined, summarize results from all analyses undertaken.*** |  |
|  |  |  | ***(c) If detailed results are available elsewhere, state how they can be accessed.*** |  |
| **Discussion** |  |  |  |  |
| *Key results* | 18 | Summarize key results with reference to study objectives. |  | The first paragraph of the discussion on page 13 summarises the key results |
| *Limitations* | 19 | Discuss limitations of the study, taking into account sources of potential bias or imprecision. Discuss both direction and magnitude of any potential bias. |  | The limitations in this study are highlighted on the last paragraph of page 15 |
| *Interpretation* | 20 | Give a cautious overall interpretation of results considering objectives, limitations, multiplicity of analyses, results from similar studies, and other relevant evidence. |  | Pages 13,14,15 in the discussion we interpret our key findings giving evidence from literature |
| *Generalizability* | 21 | Discuss the generalizability (external validity) of the study results. |  | In the final paragraph of our discussion on page 16 we mention that our study highlights the usefulness of employing diverse measures of obesity in assessing the link between obesity and cancer |
| **Other information** |  |  |  |  |
| *Funding* | 22 | Give the source of funding and the role of the funders for the present study and, if applicable, for the original study on which the present article is based. |  | On page 2 we outline all sources of funding |

STROBE: STtrengthening the Reporting of Observational Studies in Epidemiology

*Give information separately for cases and controls in case–control studies and, if applicable, for exposed and unexposed groups in cohort and cross-sectional studies.
